# Differentiating COVID-19 and dengue from other febrile illnesses in co-epidemics: Development and internal validation of COVIDENGUE scores

**DOI:** 10.1101/2021.10.14.21264897

**Authors:** Patrick Gérardin, Olivier Maillard, Léa Bruneau, Frédéric Accot, Florian Legrand, Patrice Poubeau, Rodolphe Manaquin, Fanny Andry, Antoine Bertolotti, Cécile Levin

## Abstract

**Background:** From a cohort study, we developed two scores to discriminate coronavirus 2019 (COVID-19) from dengue and other febrile illnesses (OFIs).

**Methods:** All subjects suspected of COVID-19 who attended the SARS-CoV-2 testing center of Saint-Pierre hospital, Reunion, between March 23 and May 10, 2020, were assessed for identifying predictors of both infectious diseases from a multinomial logistic regression model. Two scores were developed after weighting the odd ratios then validated by bootstrapping.

**Results:** Over 49 days, 80 COVID-19, 60 non-severe dengue and 872 OFIs were diagnosed. The translation of the best fit model yielded two scores composed of 11 criteria: contact with a COVID-19 positive case (+3 points for COVID-19; 0 point for dengue), return from travel abroad within 15 days (+3/-1), previous individual episode of dengue (+1/+3), active smoking (−3/0), body ache (0/+5), cough (0/-2), upper respiratory tract infection symptoms (−1/-1), anosmia (+7/-1), headache (0/+5), retro-orbital pain (−1/+5), and delayed presentation (>3 days) to hospital (+1/0). The area under the receiver operating characteristic curve was 0.79 (95%CI 0.76-0.82) for COVID-19 score and 0.88 (95%CI 0.85-0.90) for dengue score. Calibration was satisfactory for COVID-19 score and excellent for dengue score. For predicting COVID-19, sensitivity was 97% at the 0-point cut-off and specificity 99% at the 10-point cut-off. For predicting dengue, sensitivity was 97% at the 3-point cut-off and specificity 98% at the 11-point cut-off.

**Conclusions:** The COVIDENGUE scores proved discriminant to differentiate COVID-19 and dengue from OFIs in the context of SARS-CoV-2 testing center during a co-epidemic.

## Background

Prompt identification and management of overlapping COVID-19 and dengue infections may prevent cases of both viral diseases from deteriorating [1]. Moreover, it can avoid events of hospitalization in the Intensive Care Unit (ICU) and nosocomial transmission of both infections which may be useful whenever syndemics stress healthcare systems [2]. For public health, more rapid quarantine, contact tracing and vector control measures may help to mitigate the dynamics of both epidemics [2,3].

Since Severe Acute Respiratory Syndrome Coronavirus-2 (SARS-CoV-2) spread globally, several countries have been facing dengue epidemics with the fear of increased mortality in the most vulnerable populations [4]. Moreover, the absence of both highly sensitive and specific rapid diagnostics tests may have hampered discrimination between the two separate diagnoses possibly leading to misdiagnoses and the implementation of inadequate countermeasures in emergency situations [4]. On Reunion Island, a French overseas territory located in the southwestern Indian ocean region known to have host one of the largest chikungunya outbreaks [5], dengue virus (DENV) is endemic with annual epidemics occurring since 2015 [6]. Moreover COVID-19, which emerged in March 2020, has established an autochthonous transmission since August 2020 [7].

Surprisingly, most prediction models in the field of COVID-19 research have been dedicated to prognosis and not to the identification of people diseased from infection nor at risk of being infected [8]. Given that differential diagnosis between COVID-19 and dengue was difficult, we set-up a cohort study and developed a multinomial logistic regression model (MLR) aimed at distinguishing SARS-CoV-2 or DENV infections from other febrile illnesses (OFIs) during the first COVID-19 introductive pandemic wave [9].

Herein, we furthered our previous reflection to improve the predictive capability of our model in testing the hypothesis that the more variables included in the model, the better the discrimination between the diseases [10].

The objective of this study was to develop and internally validate a scoring system able to predict both infectious diseases which to date had never been performed.

## Methods

### Study design, study setting and population

A retrospective cohort study was conducted using prospectively collected data between 23 March and 10 May 2020, on all participants screened for COVID-19 within the UDACS (*Unité de Dépistage Ambulatoire du COVID-19 Sud*) of Saint-Pierre which is one of the two SARS-CoV-2 testing centers of the *Centre Hospitalier Universitaire Réunion* (CHU Reunion) providing care for the general population at the time. People without symptoms or with co-infections were excluded from the study.

Consecutively arriving outpatients to the SARS-CoV-2 testing center were informed of the study both verbally and by means of an informational document. Adults, as well as children under the age of 18 years (having the additional verbal consent of their parent or legal guardian) who expressed no opposition were asked to answer a questionnaire and were personally interviewed by a nurse in accordance with the French legislation on bioethics for retrospective research.

Patients’ medical records were retrospectively reviewed and anonymized data were collected in standardized forms according to the MR-004 procedure of the *Commission Nationale de l’Informatique et des Libertés* (the French Information Protection Commission). The ethical character of this study on previously collected data was approved by the Scientific Committee for COVID-19 research of CHU Reunion and anonymized data were registered on the Health Data Hub (N° F20201021104344/October, 2020).

### Data collection and gold standard procedure

The questionnaire included information on: i) demographics (gender and age), ii) occupation, iii) risk factors (smoking, obesity, return from travel abroad during the 15 previous days), iv) comorbidities (diabetes, hypertension, cardiovascular disease, chronic obstructive pulmonary disease, cancer, previous episode of dengue and “other comorbidities”), v) intra-household and individual exposure to SARS-CoV-2, vi) individual symptoms (fever, cough, dyspnea/shortness of breath, body aches, diarrhea, gut symptoms, ageusia, metallic taste, anosmia, fatigue, headache, retro-orbital pain and upper respiratory tract infection symptoms) and vii) treatment (antihypertensive drugs and/or hydroxychloroquine). Patients’ temperature, pulse rate, respiratory rate, and oxygen saturation (SpO_2_) were also measured during the consultation, as well as the presence of cough and/or anxiety. People reporting symptoms were examined by a medical resident or a senior infectious disease specialist in accordance with routine care procedures.

All participants were screened for COVID-19 by a nurse using a nasopharyngeal swab for 20 seconds in one nostril [11]. Each sample was administered for a SARS-CoV-2 reverse transcription-polymerase chain reaction (RT-PCR) using the Allplex 2019-nCov™ assay (Seegene, Seoul, Republic of Korea) or an in-house kit (CNR Pasteur) targeting N, RdRP and E genes, or N and IP2/IP4 targets of RdRP depending on which assay was used. Moreover, each participant of the study that was suspected of having dengue was tested for NS1 antigen using an OnSite™ Duo dengue Ag-IgG-IgM rapid diagnostic test (CTK Biotech, San Diego, CA, USA). If these patients had a negative result, they were explored further with a DENV RT-PCR or a dengue serology panel according to the timing of symptoms.

### Statistical analysis

Proportions between COVID-19, dengue, and non-COVID-19 non-dengue OFI patients were compared using Chi square or Fisher exact tests, where deemed appropriate. Bivariable and multivariable multinomial logistic regression (MLR) models were fitted within Stata14® (Statacorp, College Station, Texas, USA) to identify independent predictors of COVID-19 and dengue using OFIs as controls.

The first step of the process included fitting a full MLR model with all significant covariates identified by bivariable analysis [9]. From the candidate predictors, we used a backward stepwise selection procedure to drop out non-significant variables (output if *P*>0.05). At this second step, we built a parsimonious MLR model with all significant predictors. In this model, an adaption of the Hosmer-Lemeshow goodness-of-fit chi2 test was used for MLR models with polytomous categorical outcomes [12] (hereafter named MHL test for multinomial Hosmer-Lemeshow) to minimize the discrepancy between predicted and observed events. In these analyses, crude, and adjusted odds ratios (OR) and 95% confidence intervals (95% CI) were assessed using the binomial and Cornfield methods respectively.

Based on the assumption that if there are more variables, there will be better discrimination [10], we added variables that were ruled out at the borderline of significance during the backward stepwise elimination process to our previous 9-covariate parsimonious model [9].

The strategy of this third step is detailed in the text file S1.

Weighted analyses on the overall inverse probability of hospitalization to assess the potential for selection bias and to test the robustness of the identified predictors were performed next.

Lastly, from the best fit MLR compromise model, we derived two simple scores, the COVIDENGUE scores (one for COVID-19 and, one for dengue) after weighting the OR according to a predefined rule (S1 Table). This rule gave a weight to all the model covariates (no matter their significance) to maximize the possible combinations and to provide the largest range of values which, theoretically, enables the best discrimination.

The discriminative ability of the models and of the COVIDENGUE scores (*i*.*e*., the model and score performances) for the diagnosis of COVID-19 and dengue were tested using receiver operating characteristic (ROC) plot analyses which considers ROC plots and areas under ROC curves (AUC) with their 95%CIs [13]. Discrimination is usually considered as null when the AUC is 0.5, poor when between 0.5 and 0.7, satisfactory between 0.7 and 0.8, good between 0.8 and 0.9, excellent between 0.9 and 1, and perfect when the AUC equals 1. In addition, we provided classification plots to assess the discriminative ability of COVIDENGUE scores conditional to absolute risk thresholds [14]. Finally, scores performances were internally validated by using bootstrapping (2000 replicates).

The calibration of the COVIDENGUE scores (*i*.*e*., the adequacy between predicted and observed events) was evaluated using state-of-art calibration plots [15] and Hosmer-Lemeshow tests for MLR and binomial logistic regression models [12,16], as well as with event-based or risk-based calibration plots which were displayed over the range of MHL deciles of predicted risks.

The diagnostic performance of each COVIDENGUE score cut-off was displayed in terms of sensitivity, specificity, positive likelihood ratio (LR+), negative likelihood ratio (LR-), positive predictive value (PPV), negative predictive value (NPV) and diagnostic accuracy.

For all these analyses, observations with missing data were ruled out, tests were two-tailed, and a *P*-value less than 0.05 was considered as statistically significant.

The full details of the methods can be found in the text file S1. The results were reported following both the STROBE and TRIPOD reporting guidelines for observational studies and prediction models (text file S2), respectively [17,18].

## Results

### Characteristics of the study population

Between 23 March and 10 May 2020, 1,715 subjects were admitted to the UDACS for screening for or diagnosis of COVID-19. Over this 6-week period, the lab did not diagnose any cocirculation of influenza or non-influenza respiratory viruses.

As part of an expanded screening week dedicated to all admissions to our hospital, 370 subjects who were screened opportunistically and 332 fully asymptomatic subjects were ruled out leaving 1,013 outpatients eligible for this analysis. The study population is shown in Figure 1. The characteristics of the 1,013 of patients who consulted the COVID-19 screening center during the COVID-19 dengue co-epidemics are presented in S2 Table.

**Figure 1.**
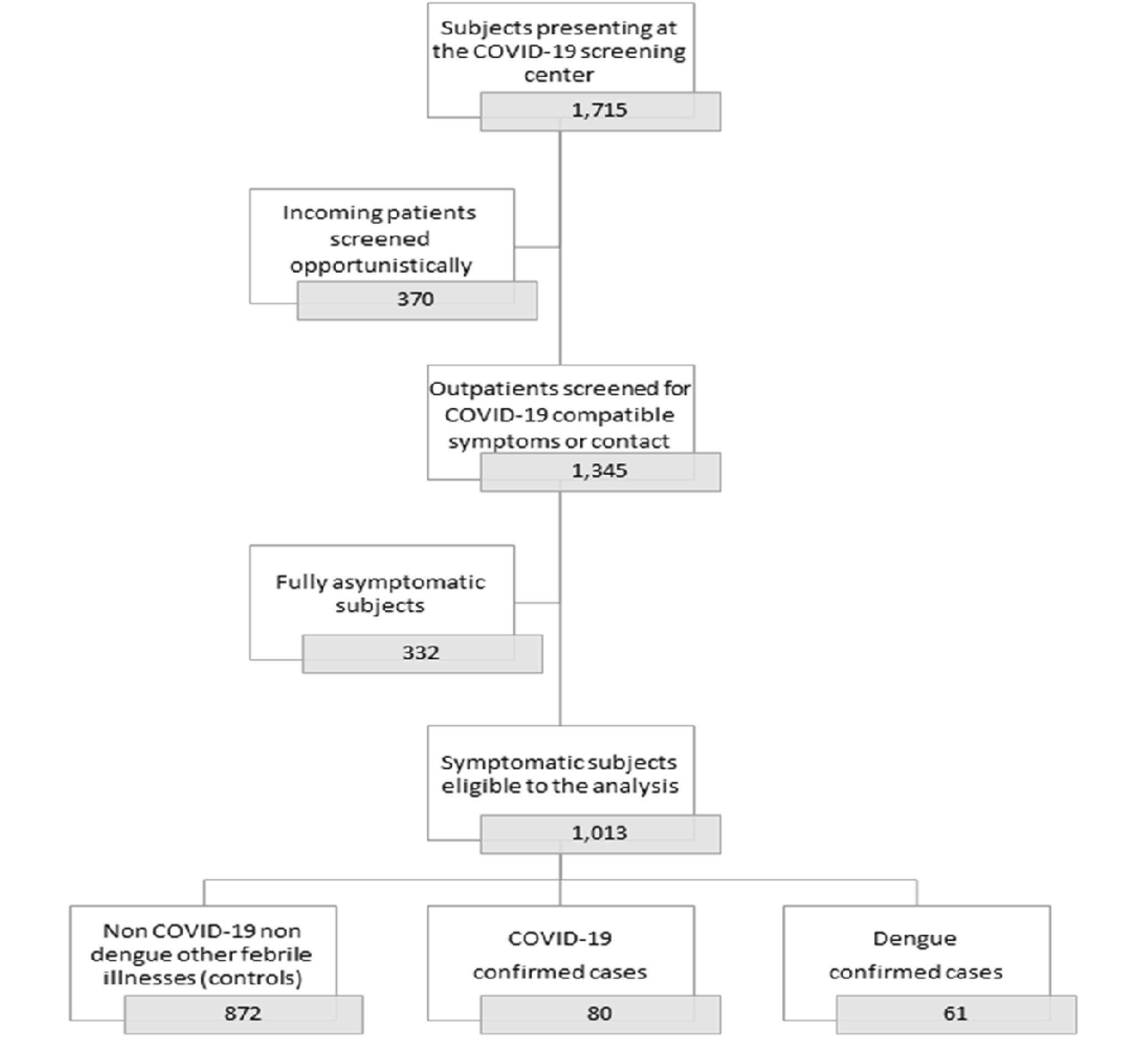
Study population for differentiating dengue and COVID-19 from other febrile illnesses in co-epidemics

COVID-19 was diagnosed in 80 patients (7.9%) and dengue in 61 patients (6.0%) while 872 patients tested negative and were clinically considered as non-COVID-19, non-dengue subjects. Interestingly, COVID-19 patients presented themselves later in the course of the illness compared to patients with dengue or OFIs (time elapsed since symptom onset, 7.5 days versus 4.2 days or 6.3 days, *P*<0.001). Dengue was more likely to be symptomatic than COVID-19 as exemplified by higher frequencies of fever, body ache, gut symptoms, fatigue, headache, and retro-orbital pain, as well as a higher need for physical examination. However, COVID-19 exhibited longer durations of fatigue and rhinorrhea.

### Bivariate and multivariate analyses

The crude relationships between the sociodemographic, epidemiological, and clinical characteristics in cases of COVID-19, dengue and OFIs are presented in S3 Table.

Bivariable analysis identified anosmia, the return from travel abroad during the previous 15 days, contact with a COVID-19 positive case and delayed presentation (beyond three days since symptom onset) as candidate predictors for COVID-19, whereas healthcare workers and active smokers as those protected against COVID-19. Headache, retro-orbital pain, body ache, fatigue, gut symptoms, and previous individual episode of dengue were potential predictors for dengue, while the recent return from travel abroad and cough were potential protective factors against dengue. Interestingly, Upper Respiratory Tract Infection (URTI) symptoms provided protection against both infections (which means they were associated with OFIs), while the role of fever and ageusia was less clear, these being associated with both diagnoses of interest (S3 Table).

The variables that were significant in the bivariable analysis were entered into a full multivariable MLR model (S4 Table). This generated 4% of missing observations (n=46). It supported the role of anosmia, contact with a COVID-19 positive case and recent return from travel abroad as independent predictors of COVID-19 as well as an association of active smokers to protection against COVID-19. In turn, previous individual episodes of dengue, body ache, headache and retro-orbital pain were independent predictors of dengue, while cough was less likely to be observed with this infection. As for the bivariable analysis, URTI symptoms were indicative of OFIs. Alternatively, fever proved non discriminant even though it was far more common with dengue which motivated its exclusion from further analyses. As in our previous analysis [9], these findings were contrasted by the weighting on the inverse probability of hospitalization rates (S5 Table).

After multicollinearity analysis, control of overfitting and unnecessary adjustments, the best fit MLR compromise model (based on AIC and BIC metrics) included eleven covariates (Table 1). This confirmed the role of anosmia, contact with a COVID-19 positive case and/or recent return from travel abroad as independent predictors of COVID-19, as well as the role of body ache, headache, previous individual episode of dengue and retro-orbital pain as predictors of dengue. Conversely, in this model, active smoking, cough, and URTI symptoms were considered as protective factors against COVID-19, dengue, or both.

**Table 1.** Final best fit multinomial logistic regression model distinguishing the independent predictors of COVID-19 and dengue from those of other febrile illnesses among 969 participants who consulted at a COVID-19 screening center in Saint Pierre (Reunion Island) during the COVID-19 dengue co-epidemics from 23 March to 10 May 2020

| Outcomes (versus other febrile illnesses as controls*) |  |  | COVID-19 (N = 73) |  |  | Dengue (N = 60) |  |  |  |  |
| --- | --- | --- | --- | --- | --- | --- | --- | --- | --- | --- |
| Predictors | n | CIR, (%) | aOR | 95% CI | Points | n | CIR, (%) | aOR | 95% CI | Points |
| Contact with a COVID-19 positive case | 40 | 15.38 | **4.26** | **2.33 to 7.78** | **+ 3** | 6 | 2.31 | 0.71 | 0.26 to 1.87 | 0 |
| Return from travel abroad < 15 days | 38 | 16.03 | **3.53** | **2.05 to 6.05** | **+ 3** | 6 | 2.53 | 0.41 | 0.15 to 1.05 | - 1 |
| Previous individual episode of dengue | 4 | 9.06 | 2.16 | 0.56 to 8.29 | + 1 | 9 | 20.45 | **4.50** | **1.68 to 12.00** | **+ 3** |
| Active smoker † | 4 | 2.53 | **0.25** | **0.09 to 0.67** | **- 3** | 12 | 7.59 | 1.38 | 0.65 to 2.91 | 0 |
| Cough | 32 | 6.82 | 0.95 | 0.53 to 1.68 | 0 | 17 | 3.62 | **0.36** | **0.18 to 0.70** | **- 2** |
| Body ache ‡ | 29 | 7.09 | 1.26 | 0.70 to 2.24 | 0 | 52 | 12.71 | **6.35** | **2.56 to 15.71** | **+ 5** |
| Anosmia | 26 | 27.96 | **7.64** | **4.13 to 14.12** | **+ 7** | 3 | 3.23 | 0.47 | 0.11 to 2.01 | - 1 |
| Headache | 28 | 5.70 | 0.92 | 0.54 to 1.55 | 0 | 55 | 11.20 | **5.51** | **1.85 to 16.41** | **+ 5** |
| Retro-orbital pain | 1 | 2.27 | 0.53 | 0.06 to 4.53 | - 1 | 17 | 38.64 | **4.42** | **2.03 to 9.59** | **+ 5** |
| Upper respiratory tract infection symptoms # | 28 | 5.63 | 0.53 | **0.31 to 0.90** | **- 1** | 20 | 4.02 | **0.51** | **0.26 to 0.98** | **- 1** |
| Presentation > 3 days after symptom onset | 53 | 9.55 | 1.71 | 0.94 to 3.10 | + 1 | 24 | 4.32 | 0.75 | 0.39 to 1.42 | 0 |

Goodness-of-fit and discrimination indicators of the MLR models are displayed in S6 Table. We made the assumption that the best fit compromise model could be achieved with the eleven abovementioned covariates. The AUCs of the model were of 0.80 (95%CI 0.74-0.86) for COVID-19 and 0.89 (95%CI 0.84-0.92) for dengue in the primary analysis (Figure 2, panel a), and of 0.80 (95%CI 0.73-0.86) and 0.88 (95%CI 0.84-0.92) after bootstrapping, respectively. Further adjustment on both age and/or gender did not improve the AUC of the model to a point sufficient enough to change interpretation when considering AUC boundaries (S6 Table).

**Figure 2.**
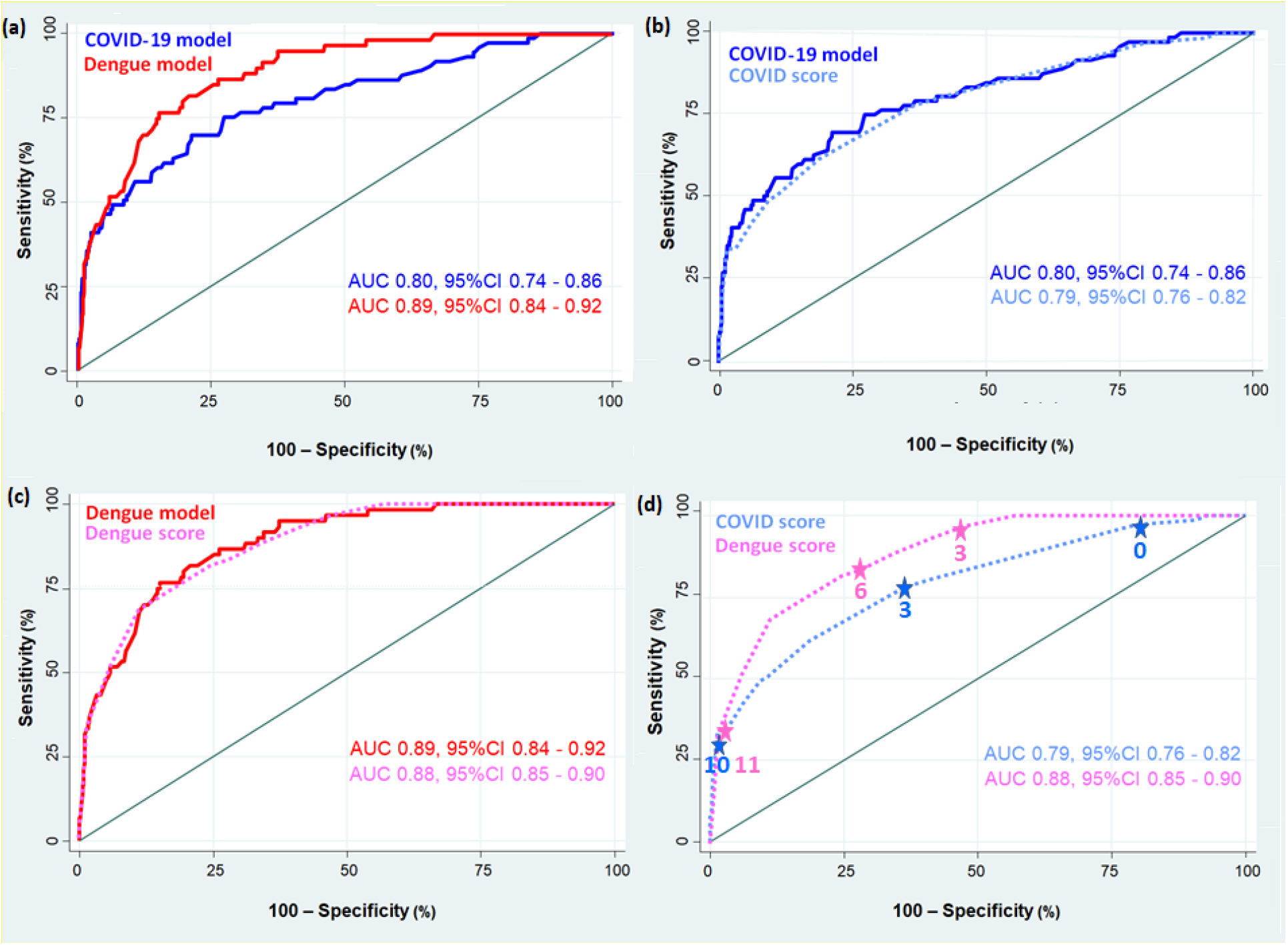
Discriminative ability of the final model and of COVIDENGUE scores for COVID-19 and dengue at Saint-Pierre, Reunion Island from 23 March to 10 May 2020 Notes: Areas under receiver operating characteristic (AUC) curves are given for the best fit compromise final models (reference models) and the COVIDENGUE scores before validation by bootstrapping. The better the model or the score discriminates, the more the ROC curve approaches the upper left corner of the plot. A model with no discriminative ability has a true ROC curve that lies on the diagonal line.

### Score development and internal validation

Based on this final model, we derived two COVIDENGUE scores. For a better appropriation, we decided to set the weight of each dengue clinical criterion at 5 points. With this exception, the weighting scale was as described in S1 Table.

The translation of the final MLR model into two COVIDENGUE scores was satisfactory. This step showed barely a 1-point loss in the discriminative ability for both COVID-19 and dengue and a superimposition of model and score ROC plots (Figure 2, panel b and panel c).

The median value of the COVID-19 score was 1 (Q_1_-Q_3_: 0 to 3, range: −4 to 14) and the median value of the dengue score was 3 (Q_1_-Q_3_: −1 to 7, range: −4 to 18). In the primary analysis, the discriminative ability of the scores was satisfactory to good for COVID-19 (AUC 0.79, 95%CI 0.76-0.82) and good for dengue (AUC 0.88, 95%CI 0.85-0.90) (Figure 2, panel d). Interestingly, the dengue score exhibited higher true positive rates than the COVID-19 score (which means false negative cost was minimized versus the false positive cost) in risk thresholds higher than 0.4. COVID-19 had higher false positive rates in risk thresholds under 0.5 than the dengue scores (which means FP cost was minimized versus FN) (S1 Fig.).

After bootstrapping, when considering confidence intervals, the discriminative ability was between poor and good for COVID-19 (AUC 0.75, 95%CI 0.68-0.82) and good for dengue (AUC 0.86, 95%CI 0.81-0.90), which represented an AUC loss of five points and three points from the final MLR model, respectively.

Overall, the calibration of the COVIDENGUE scores as shown by calibration plots (S1 Fig.) and MHL tests (S7 Table and S8 Table) was deemed satisfactory albeit prone to underfitting for COVID-19 (slope: 1.22, intercept 0.08; chi2 (6) 1.75, *P*=0.9416), whereas it was excellent and consensual for dengue (slope: 0.97, intercept 0.12; chi2 (8) 6.78, *P*=0.5605). Strikingly, despite few discrepancies between predicted and observed events (S2 Fig. and S3 Fig., panel a and panel c), and a trend towards underprediction of COVID-19 events (S4 Fig., panel a), the MHL test of both COVIDENGUE scores was well balanced across the deciles of predicted risks (S7 Table and S8 Table) and their calibration displayed other metrics better than those of the reference model (S9 Table).

For overall COVID-19 prediction, a threshold of 0 points displayed a sensitivity of 97.3% (95%CI 90.5%-99.7%) and a NPV of 99.0% (95%CI 96.3%-99.8%), while a threshold of 10 points displayed a specificity of 98.8% (95%CI 97.8%-99.4%) and a PPV of 65.1% (95%CI 48.6%-78.6%). A 3-point cut-off maximized both sensitivity and specificity (78.1% and 63.7%, respectively). Regardless of COVID-19 prevalence, the COVID-19 score was effective in excluding COVID-19 (LR-<0.10) for negative score values under −1, whereas it was effective for diagnosing COVID-19 (LR+ >10) with score values higher than 7. The detailed performances of the COVID-19 score are presented in S10 Table, its intrinsic and extrinsic properties in Figure 3 (Panel a to panel d). For COVID-19 individual risk prediction, the estimated probability of being infected derived from the COVID-19 score was Prob_(Y=1/x)_= 1 / (1 + exp^-(−3.698086 + 0.3533028 X score value)^).

**Figure 3.**
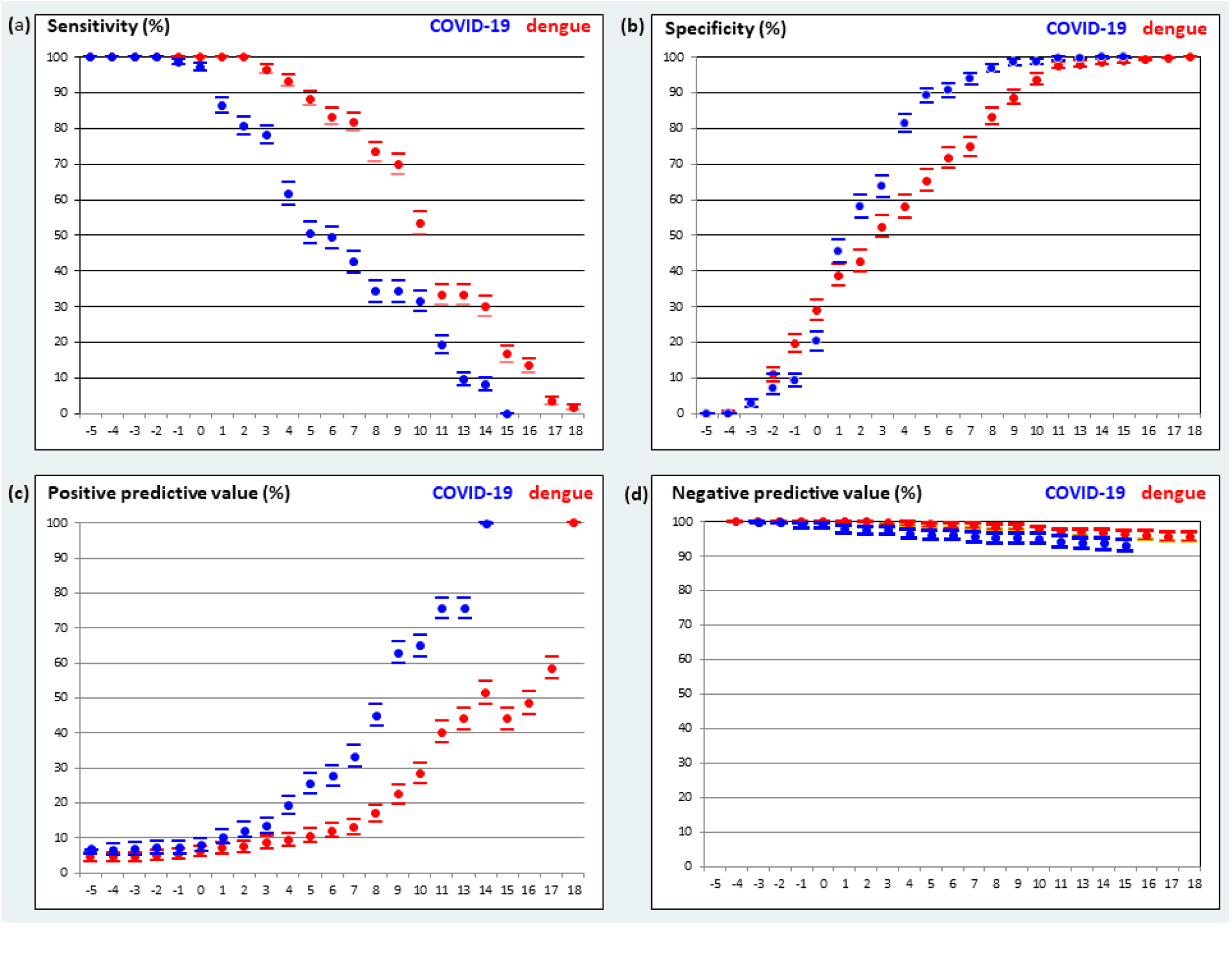
Diagnostic performances of the COVIDENGUE scores

For dengue overall prediction, a threshold of 3 points displayed a sensitivity of 96.7% (95%CI 88.5%-99.6%) and a NPV of 99.7% (95%CI 98.8%-99.9%) while a threshold of 11 points displayed a specificity of 97.7% (95%CI 96.5%-98.6%) and PPV of 40.3% (95%CI 27.9%-54.0%). A 9-point cut-off maximized both sensitivity and specificity (70.0% and 88.7%, respectively). Regardless of dengue prevalence, the dengue score was effective in excluding dengue (LR-<0.10) for score values under 4, whereas it was effective for diagnosing dengue (LR+ >10) with score values higher than 10. The detailed performances of the dengue score are presented in S11 Table, its intrinsic and extrinsic properties in Figure 3 (Panel a to panel d). For dengue individual risk prediction, the estimated probability of being infected derived from the dengue score was Prob_(Y=1/x)_ = 1 / (1 + exp^-(−5.508826 + 0.4406125 X score value)^).

## Discussion

In this study, COVIDENGUE scores were developed and internally validated. An eleven covariate-based prediction model aimed at distinguishing COVID-19 from dengue and other febrile illnesses at their clinical onset was made in the context of cocirculation of SARS-CoV-2 and DENV. Overall, the predictive performance of the score, meaning the ability to diagnose or to rule out infection, was good for dengue and satisfactory for COVID-19, while calibration performance meaning the ability to minimize the discrepancy between expected and observed events, was at least satisfactory on multiple metrics of goodness-of-fit statistics. The dengue score for which symptoms were good predictors, displayed both better sensitivity and a higher negative predictive value while the COVID-19 score for which risk factors were paramount, proved more specific and had a higher positive predictive value across the whole range of cut-offs.

### Strengths and limitations

A scoring system should have both construct and content validity. It must be able to be reproduced over time and across geographic and methodological boundaries. It must also be accurate (calibration and discrimination) and clinically meaningful [19]. Before this study, we previously assumed that COVID-19 and dengue diagnoses could be affected by a misclassification bias which could stem from the poor sensitivity of both SARS-CoV-2 molecular and DENV NS1 antigen tests rather than from their false positives [9]. This putative bias was believed to be minimal given that, firstly, on Reunion Island, like anywhere else during the rise of the COVID-19 pandemic [20], there was little cocirculation of other respiratory viruses that could have competed with SARS-CoV-2 and caused false negatives, [21] and secondly, for COVID-19, negative samples were retested by RT-PCR upon onset of new symptoms, meaning that rapid antibody or antigenic tests were ruled out, while for dengue, the workup was completed by a RT-PCR or a serology test to downsize false negative and false positive proportions [22-24]. This caution decision rule likely pledges the diagnostic accuracy of our gold standards.

Interestingly, the calibration of both scores was satisfactory to excellent and displayed measures close to the theoretical model from which they were derived based on a set of various goodness-of-fit metrics. Notwithstanding a relatively small study population, the diagnostic accuracy of gold standards, along with the acceptable calibration properties ensure the validity of construct of the COVIDENGUE scores and the reliability of their predictions at the individual level for the dengue score, while, together with their discriminative ability, also lend support to their clinical utility [25].

The COVIDENGUE scores were developed using MLR [9] which is the gold standard method for assessing non-ordered polytomous categorical outcomes [26,27]. Except for active smoking whose protective effect against SARS-CoV-2 infection remains a matter of debate among researchers [28,29], all predictors retained to build the scores had been previously identified as relevant indicators of COVID-19 or dengue or associated with OFIs (especially respiratory infections) [30-42] This ensures both the validity of content of this scoring system and the possibility of contrasted predictions. For example, international travel had been identified as a source of COVID-19 during the first pandemic wave [35-37]. In a Colombian study, dengue proved more symptomatic than COVID-19 and dengue patients came to the hospital in greater numbers than COVID-19 patients [38]. In an attempt to differentiate COVID-19 from influenza or from dengue based on three distinct Singaporean cohorts, upper respiratory tract symptoms pointed to influenza, while headache and joint pain pointed to dengue, as in our study [39]. In Brazil’s Amazonian basin, a previous dengue episode, as diagnosed by positive IgG antibodies, was associated with twice the risk of clinically apparent COVID-19 [40]. The external validity of the other predictors has been discussed thoroughly in our previous study [9]. Interestingly, similar to our first analysis, weighting on the inverse odds of hospitalization abrogated the significance of a few predictors (Table 1 *versus* S5 Table), which suggests a contrast in our findings at the population level and motivates further validation studies in the primary care setting.

Notably, this hospital-based study was conducted in a SARS-CoV-2 screening center which may have underestimated the real incidence of dengue and introduced another information bias in that dengue patients could have been potentially directed towards other units or even underreported given the lack of epidemiological predictors reported for dengue [9]. This potential limitation of the validity of content should be investigated in future studies by adding more risk factors for dengue to refine our models. Lastly, our scoring system was composed of only clinical and epidemiological criteria. It was user-friendly for diagnosis purposes which should facilitate its utilization across different settings while also helping its external validation.

### Interpretation

For epidemiological and clinical practices, the overall performance of a prediction model relies first on its discrimination [13]. In this perspective, ROC plots do not offer more information than the AUC to indicate the discriminative ability [13,14]. In this study, we demonstrated that the discriminative ability of the models could be improved only at the unreasonable cost of complexity (20-item model), or when adding age to the COVID-19 model; a factor whose effect might change according to the context (S6 Table). This study also showed that the translation of the model into two scores was not accompanied by a significant loss in discriminative ability (Figure 2) which suggests an adequate weighting of the scores.

Importantly, we provided classification plots which may offer more information for decision-making conditional to risk thresholds [13,14]. Overall, classification plots may reveal a better ability of the COVID-19 score to predict non-events (OFIs) and a better ability of dengue score to predict events (dengue). Moreover, at lower risk thresholds the COVID-19 score exhibited a lower cost of FP than the dengue score. For example, when the event risk was 0.2, the COVID-19 score yielded a 0.5:1 FN to FP ratio while it was 1:1 for the dengue score (Fig. S1). These results aligned with a trend towards a better specificity for the COVID-19 score than for the dengue score across the range of cut-off values (Figure 3). Conversely, at higher risk thresholds (>0.5), when it came to predict an event, the dengue score displayed a better sensitivity (in other words, higher TP rate, or a lower cost of FN *versus* FP cost). Taken together with respect to the SARS-CoV-2 strategy of testing, isolating, and tracing, our findings encourage evaluating the addition of clinical or biological discriminative variables [40,41] in the COVID-19 score to improve its sensitivity across the risk thresholds while in regard to the dengue strategy of testing, isolating, and targeted vector control, they encourage the fitting with more specific epidemiological variables highly predictive of an infective bite by an *Aedes* mosquito.

At an individual risk level, predictions should be guided using first of all calibration performances (calibration plots and goodness-of-fit metrics) [24]. Our findings showed that the calibration of the models (Table S6 to S8) and their derived scores (Table S9) were excellent and consensual for dengue which enables individual risk prediction and satisfactory for COVID-19 which suggests caution for individual risk prediction.

### Generalizability

The COVIDENGUE scores were developed from data acquired within a hospital-based SARS-CoV-2 testing center on Reunion Island which is a tropical setting where dengue co-circulated early on during the first pandemic wave at a time when there was no possibility to screen for COVID-19 outside the hospital. The circulation of the SARS-CoV-2 variant was furthermore unknown and the population of infected people were mainly composed of relatively healthy travelers [9,36,37]. Thus, although our center served an ambulatory healthcare driven population, the scores will have to be validated in primary care settings before being broadly used in the community. They will also have to be validated in the highly comorbid autochthonous population of the island [9,36,37].

In future research, the scores should be studied in the context of newly circulating SARS-CoV-2 variants as well as in the context of populations immunized against dengue or COVID-19. On Reunion Island, the first wave of SARS-CoV-2 circulation ended in June 2020 as a result of the influence of the first national lockdown. The second wave began in August 2020 during the winter season of the region and was concomitant to the spread of the D614G mutation in Europe [7]. Since this period, SARS-CoV-2 transmission has been mainly autochthonous and successively maintained through the circulation of both South African (B.1.351/501Y.V2) and the Indian (B.1.617/21APR-02) variants of concerns (VOCs). While the clinical presentation of dengue appears to be different between DENV-2 and DENV-1 serotype infections (DENV-1 has been predominant on the island since 2020) on top of the proportions of secondary infections [43], it is not yet clear whether the prevalence of COVID-19 has changed throughout the circulating VOCs despite a trend towards increased severity reported with the UK (B.1.1.7/501Y.V1) [44], along with the Brazilian (P.1/501Y.V3) [45], South African [46], or Indian variants [47]. The same could be said to the potential for more clinically apparent manifestations of COVID-19 when DENV infection precedes SARS-CoV-2 infection [40] as well as the potential for higher severity with SARS-CoV-2 DENV co-infections [2]. Both have to be fully investigated in the future with the diagnostic value of the COVIDENGUE scores evaluated.

### Implications

For individual risk prediction and clinical practice, the equations of estimated probability of being diagnosed as infected derived from the scores could be used to define individual risks conditional to adequate calibration. Herein, we have shown that the COVIDENGUE scores could be useful to diagnose dengue patients in a tropical SARS-CoV-2 screening center, however they deserved further improvements for diagnosing COVID-19.

For public health purposes, the score values could be incorporated into testing strategies and guided with mitigation interventions whenever routine biological testing is ineffective. For clinical research, the cut-off values could serve to risk stratification in new diagnostic studies and the COVIDENGUE scores items incorporated into propensity scores. For benchmarking of prediction models, the predicted risk probabilities of a new model could also be summed up and compared to the total number of infected individuals (to define risk thresholds) under the assumption that such an observed-to-predicted risk ratio together with the slope of the calibration plots and the Hosmer-Lemeshow chi2 test probability, would be close to one, to underlie the adequate calibration of the new model.

When it comes to testing the external validity of the COVIDENGUE scores in a different epidemiological context, or to improving their predictive performances by adding or removing variables within a new model, investigators will have to consider using calibration plots and goodness-of-fit statistics to see whether the model is properly calibrated and could apply to individual risk prediction. Such novel investigations should delve deeper into demonstrating the clinical use while providing new indicators such as the IDI (Integrated Discrimination Improvement) and/or the net benefit from classification plots or decision curve analyses [13,14].

### Conclusions

In conclusion, the COVIDENGUE scores proved discriminant to differentiate COVID-19 and dengue from OFIs in the context of SARS-CoV-2 testing center during a co-epidemic. Further studies are needed to validate or refine these scores in other settings.

## Supporting information

Methodological appendix

STROBE and TRIPOD checklists

## Data Availability

Data are entirely accessible through a data repository through the publication of this article.

https://doi.org/10.1371/journal.pntd.0008879.s007

## Acknowledgments

The authors are indebted to the staffs of the Department of Infectious Diseases and Tropical Medicine, especially Dr Yatrika Koumar, and the SARS-CoV-2 testing center with special appreciation given to Dr Antoine Joubert and the nurses who performed the questionnaires. The authors would also like to thank the biologists at CHU de La Réunion for their timely diagnosis, the participants for their interest in research and AcaciaTools for their reviewing and medical writing services.

## Supporting information

Supplementary material is available online. While the supplementary tables have been copyedited, the methodological appendix and the Venn diagram have not been copyedited and are the sole responsibility of the authors. Questions and comments about these should be addressed to the corresponding author. STROBE and TRIPOD checklists (text file S2).

## Notes

### Author’s contributions

Conceptualization: Fanny Andry, Patrick Gérardin, Cécile Levin. Data curation: Patrick Gérardin. Formal analysis: Patrick Gérardin, Antoine Bertolotti, Olivier Maillard, Léa Bruneau, Cécile Levin. Investigation: Frédéric Accot, Florian Legrand, Patrice Poubeau, Rodolphe Manaquin, Fanny Andry, Antoine Bertolotti, Cécile Levin. Methodology: Patrick Gérardin. Project administration: Fanny Andry. Resources: Cécile Levin. Software: Patrick Gérardin. Supervision: Olivier Maillard, Antoine Bertolotti. Validation: Patrick Gérardin, Cécile Levin. Visualization: Patrick Gérardin. Writing – original draft: Patrick Gérardin. Writing – review & editing: Patrick Gérardin, Olivier Maillard, Léa Bruneau, Frédéric Accot, Florian Legrand, Patrice Poubeau, Rodolphe Manaquin, Fanny Andry, Antoine Bertolotti and Cécile Levin.

#### List of abbreviations (alphabetic order)

aOR: Adjusted odds ratio
AUC: Area under receiver operating characteristic curve
CIR: Cumulative incidence rate (attack rate)
CNR: *Centre national de reference*
COVID-19: Coronavirus 2019
DENV: Dengue virus
FN: False negative
FP: False positive
ICU: Intensive Care Unit
LR-: Negative likelihood ratio
LR+: Positive likelihood ratio
MHL test: Multinomial Hosmer-Lemeshow test
MLR: Multinomial logistic regression
NPV: Negative predictive value
OFIs: Other febrile illnesses
PPV: Positive predictive value
RT-PCR: Reverse transcription – polymerase chain reaction
SARS-CoV-2: Severe Acute Respiratory Syndrome Coronavirus 2
SpO2: Pulse oxymetry
TN: True negative
TP: True positive
UDACS: *Unite de dépistage ambulatoire du coronavirus sud*
URTI: Upper respiratory tract infection
VOCs: variants of concerns
95% CI: 95% confidence interval

## Data availability statement

Data are entirely accessible via a data repository through the publication of this article.

