## Supplementary material for "Differentiating COVID-19 and dengue from other febrile illnesses in co-epidemics: Development and internal validation of COVIDENGUE scores": Methodological appendix

**S1 text file. Detailed Methods**

***Study design, setting and population***

A retrospective cohort study was conducted using data collected prospectively between 23 March and 10 May 2020. All subjects were screened for COVID-19 within the UDACS of Saint-Pierre (*Unité de Dépistage Ambulatoire du COVID-19 Sud*) which is one of the two SARS-CoV-2 testing centers of the *Centre Hospitalier Universitaire* (CHU) in Reunion Island serving the general population at that time [1]. It is worth noting that when SARS-CoV-2 first emerged on the island in March 2020, a dengue epidemic was already expanding. The UDACS of Saint-Pierre was mobilized as a second line for the reception system for COVID-19 patients with the frontline being the emergency units and a dedicated hospital for COVID-19 patients (CHU Félix Guyon in Saint-Denis) located near the prefecture and international airport. People without symptoms or with co-infections were excluded from the study.

***Ethics statement***

Consecutively arriving outpatients to the SARS-CoV-2 testing center were informed of the study both verbally and by means of an informational document. Adults, as well as children under 18 years old having additional verbal consent of their parent or legal guardian, who expressed their consent in participating in the study were asked to answer a questionnaire and participate in a face-to-face interview with a nurse in accordance with French legislation on bioethics for retrospective research. Patient medical records were retrospectively reviewed and anonymized data were collected using standardized forms in accordance with the MR-004 procedure of the French information protection commission*, Commission Nationale de l’Informatique et des Libertés* (CNIL)*.* The ethical character of this study, which was based on previously collected data, was approved by the Scientific Committee for COVID-19 research of the CHU Reunion and anonymized data were registered on the Health Data Hub database (N° F20201021104344/October, 2020).

***Data collection***

The items in the questionnaire included: i) demographic information (gender and age), ii) occupation, iii) risk factors (smoking, obesity and any return from travel abroad during the 15 previous days), iv) comorbidities (diabetes, hypertension, cardiovascular disease, chronic obstructive pulmonary disease, cancer, previous episode(s) of dengue and “other comorbidities”), v) intra-household and individual exposure to SARS-CoV-2 and vi) individual symptoms (fever, cough, dyspnea/shortness of breath, body aches, diarrhea, nausea, vomiting, abdominal pain, ageusia, dysgeusia/metallic taste, anosmia, fatigue, headache, retro-orbital pain and upper respiratory tract infection symptoms including sore throat, runny nose, nasal congestion and sneezing, and any treatments such as antihypertensive drugs and/or hydroxychloroquine). Patients’ temperature, pulse rate, respiratory rate, and oxygen saturation (SpO_2_) were also measured during the consultation. The presence of a cough and/or anxiety were also noted. Participants reporting symptoms were examined by a medical resident or a senior infectious disease specialist according to routine care procedures.

***Gold standard procedures***

All participants were screened for COVID-19 by a nurse using a nasopharyngeal swab for 20 seconds in one nostril [2]. Each sample was processed for a SARS-CoV-2 reverse transcription-polymerase chain reaction (RT-PCR) using a Microlab NIMBUS/STARlet IVD (Seegene, Seoul, Republic of Korea) or a MagNa Pure Compact (Roche Life Science, Penzberg, Germany) automaton for nucleic acid extraction.

SARS-CoV-2 ribonucleic acid sequences were identified using the Allplex 2019-nCov assay (Seegene, Seoul, Republic of Korea) or an in-house kit (CNR Pasteur) targeting N, RdRP and E genes, or N and IP2/IP4 targets of RdRP respectively. SARS-CoV-2 genome sequences were amplified using a Bio-Rad CFX96 (BioRad, Hercules, CA, USA) or a LightCycler 480 (Roche Diagnostics, Basel, Switzerland) system.

Patient testing negative for SARS-CoV-2 were re-tested for COVID-19 in case of new symptoms in addition to a 21-pathogen fast-track battery for respiratory viruses. Furthermore, each patient suspected of dengue was tested for NS1 antigen using an OnSite Duo dengue Ag-IgG-IgM rapid diagnostic test (CTK Biotech, San Diego, CA, USA), and if negative, tested further with a DENV RT-PCR (NucliSens easyMAG, Biomerieux, France; LightCycler 480, Roche Diagnostics, Basel, Switzerland) or a Panbio dengue Duo (IgM/IgG) capture ELISA (Abbott, Chicago, IL, USA) depending on the date of symptom onset. Patients requiring hospitalization were promptly transferred from UDACS Saint-Pierre to the COVID-19 units. Patients not requiring hospitalization were discharged to their place of residence and benefited from an ambulatory follow-up ensured by general practitioners.

***Statistical analysis***

Proportions between COVID-19, dengue and non-COVID-19 non-dengue OFI subjects were compared using Chi square or Fisher exact tests when deemed appropriate. Bivariable and multivariable multinomial logistic regression models were fitted within Stata14® (Statacorp, College Station, Texas, USA) to identify independent predictors of COVID-19 and dengue using OFIs as controls.

A full Multivariable Regression Model (MLR) was first fitted with all significant covariates identified by bivariable analysis [1]. From the candidate predictors, we used a backward stepwise selection procedure to drop out non-significant variables (output if *P*>0.05). For this second step, a parsimonious multinomial logistic regression model was built with all significant predictors. In this model, we used *mlogitgof* (an adaption of the Hosmer-Lemeshow goodness-of-fit Chi2 test (hereafter named MHL test) for MLR with polytomous categorical outcomes [3] to minimize the discrepancy between predicted and observed events. In these analyses, both crude and adjusted odds ratios (OR) and 95% confidence intervals (95% CI) were assessed using the binomial and Cornfield methods respectively [1].

Based on the assumption that if there are more variables, there will be better discrimination [4], we added variables that were ruled out at the borderline of significance during the backward stepwise elimination process to our previous 9-covariate parsimonious model [1]. This step was guided by the optimization of both the MHL test *P*-values and the Area Under Receiver Operating Characteristic curve (AUC), as well as the need for reproducibility between the different epidemiological contexts. Age and gender, whose effect directions could change conditional to the context, were therefore discarded. Moreover, based on past experience with the Chikungunya virus epidemic, we considered that the signification of these two variables may change across epidemiological settings according to occupational exposure or immune background and thus would have been inconsistent to force these into the model, which limited their interest for external validation.

For the enrichment phase of our modeling strategy, we switched to a combined approach that balanced prediction and explained using a set of ten information-based and parsimony-based metrics to identify the best fit MLR model while accounting for overfitting, underfitting, model selection uncertainty and the principle of parsimony. Under this average-ranking approach, we preselected five candidate models and rated the goodness of fit performance of each model using a 5-point scale (best fit: 1, worst: 5). The best fit MLR model for inference being the one with the lowest estimated score [5]. This rating scale included the abovementioned MHL test along with four metrics that are displayed in the *fitstat* package [6], namely the Mac Fadden’s R2, the deviance, the Akaike (AIC) and the Bayesian (BIC) information criteria. Among these standard error and prediction-based criteria, we privileged the latter two to select the best fit MLR model. The AIC and the BIC were deemed superior to the others since they maximize the calibration of the model while preventing overfitting therefore minimizing the loss of information and supporting the principle of parsimony [7,8].

Weighted analyses on the overall inverse probability of hospitalization to assess the potential for selection bias and to test the robustness of the identified predictors were performed next [1]. Briefly, we applied the hospitalization rates observed for confirmed COVID-19 and dengue cases in logistic regression models over the study period with the actual rate (1.5%) for OFIs using the *svy* function in Stata.

Lastly, from the best fit compromise model, two simple scores were derived for the COVIDENGUE scores; one was for COVID-19, and the other for dengue, after weighting the OR according to a predefined rule. This rule gave weight to all the model covariates (no matter their significance) to maximize the possible combinations and provide the largest range of values which theoretically enabled the best discrimination.

The discriminative ability of the models and of the COVIDENGUE scores (in other words, the model and score predictive performances) for the diagnosis of COVID-19 and dengue were tested using receiver operating characteristic (ROC) plot analyses [9] that consider ROC plots and areas under ROC curves (AUC) with their 95%CI (for example: the *mlogitroc* module for estimating multiclass ROC curves and AUC from multinomial logistic regression) [10]. Discrimination is typically considered as null when AUC is 0.5, poor when between 0.5 and 0.7, satisfactory between 0.7 and 0.8, good between 0.8 and 0.9, excellent between 0.9 and 1, and perfect when the AUC equals 1. AUCs were compared both graphically and by using the nonparametric DeLong method (*roccomp and rocgold* Stata commands) [11] taking either the full or ‘saturated’ main-effect model or the best fit compromise model as referent models to identify which model could both approximate the discriminatory power of the full model and be significantly more discriminant than the minimal-effect (*‘*parsimonious’) model. In addition, classification plots were provided to assess the discriminative ability of the COVIDENGUE scores conditional to absolute risk thresholds [12]. Score performances were internally validated using bootstrapping (2000 replicates).

The calibration of the COVIDENGUE scores (the adequacy between predicted and observed events) was evaluated using multiple criteria as recommended [13]. We used both state-of-art calibration plots [9,12-15], the Hosmer-Lemeshow tests for multinomial and binomial logistic regression models, with *mlogitgof and estat gof (*formerly *lfit)* commands respectively [3,16], as well as original event-based or risk-based MHL calibration plots which were displayed over the range of MHL deciles of predicted risks. Overall, we considered that ≤20% difference to the best theoretical slope and intercept values (Best slope value= 1, exponential to the best intercept value [0]= 1), or a stringent MHL test *P*-value<0.5 (which means miscalibration is defined for values <0.8 or >1.20, or a highly conservative *P*-value rejecting the alternative hypothesis; since a 20% discrepancy cut-off is often chosen in epidemiology to indicate significant changes in parameter measures) may cause potential harm to the calibration of the model and would prevent its use for individual risk prediction across the range of predicted risks.

The diagnostic performance of each COVIDENGUE score cut-off was displayed in terms of sensitivity (Sen), specificity (Spe), positive likelihood ratio (LR+), negative likelihood ratio (LR-), positive predictive value (PPV), negative predictive value (NPV) and diagnostic accuracy. Ninety-five percent confidence intervals of Sen and Spe were estimated using the exact binomial method. For LR+, LR-, PPV and NPV, 95%CIs were calculated using the maximum likelihood method assuming the prevalence was known exactly (*i.e.*, we used sample observed prevalence for PPV and NPV estimations and the substitution formula adding 0.5 to all cell frequencies when needed for LR+ and LR- calculations). The optimal cut-point was sought using Liu’s method after (2000 replicate) bootstrapping (*cutpt* module in Stata) [17]. For all these analyses, observations with missing data were ruled out, tests were two-tailed, and a *P*-value<0.05 was considered as statistically significant. This article followed both the STROBE and TRIPOD reporting guidelines for observational studies and prediction models, respectively [18,19].
