## Supplementary tables and figures for "Differentiating COVID-19 and dengue from other febrile illnesses in co-epidemics: Development and internal validation of COVIDENGUE scores"

| **S1 Table. COVIDENGUE scores weighting rules** | | | | | | | |
| --- | --- | --- | --- | --- | --- | --- | --- |
| Odds Ratio | 0.13 to 0.15 | 0.15 to 0.18 | 0.18 to 0.22 | 0.22 to 0.285 | 0.285 to 00.4 | 0.4 to 0.66 | 0.66 to 1.5 |
| Weights | -6 | -5 | -4 | -3 | -2 | -1 | 0 |
| Odds Ratio | 1.5 to 2.5 | 2.5 to 3.5 | 3.5 to 4.5 | 4.5 to 5.5 | 5.5 to 6.5 | 6.5 to 7.5 | 7.5 to 8.5 |
| Weights | 1 | 2 | 3 | 4 | 5 | 6 | 7 |
| Accordingly, weighting applied to the 11 covariates included in the COVIDENGUE scores and was as follows: i) contact with a COVID-19+ case (+3 points for COVID-19 / 0 point for dengue), ii) return from travel abroad within 15 days (+3 / -1), iii) previous individual episode of dengue (+1 / +3), iv) active smoking (-3 / 0), v) body aches (0 / +5), cough (0 / -2), vi) URTI symptoms (-1 / -1), vii) anosmia (+7 / -1), viii) headache (0 / +5), ix) retro-orbital pain (-1 / +5), and x) delayed presentation (>3 days) to hospital (+1 / 0). | | | | | | | |

| S2 Table. Characteristics of 1,013 participants who consulted a COVID-19 screening center in Saint-Pierre (Reunion Island) during the COVID-19 dengue co-epidemics from 23 March to 10 May 2020 | | | | | | | |
| --- | --- | --- | --- | --- | --- | --- | --- |
| Outcomes | **Other febrile illnesses (n = 872)** | | **COVID-19**  **(n = 80)** | | **Dengue**  **(n = 61)** | |  |
| Variables | **n** | **(%)** | **n** | **(%)** | **N** | **(%)** | ***P* value** |
| Male gender | 343 | 39.3 | 33 | 41.2 | 31 | 50.8 | 0.205 |
| Age (years), µ ± SD | 38.7 | 16.2 | 39.2 | 18.4 | 42.0 | 13.4 | 0.280 |
| 0 to 30 (Q1) | 253 | 29.0 | 28 | 35.0 | 8 | 13.1 | **0.002** |
| 31 to 41 (Q2) | 247 | 28.3 | 10 | 12.5 | 26 | 42.6 |  |
| 42 to 54 (Q3) | 221 | 25.3 | 27 | 33.7 | 13 | 21.3 |  |
| 55 to 94 (Q4) | 151 | 17.3 | 15 | 18.7 | 14 | 22.9 |  |
| Contact with a COVID-19 positive case | 231 | 26.5 | 42 | 52.5 | 6 | 9.8 | **<0.001** |
| Return from travel abroad < 15 days | 202 | 23.2 | 42 | 53.2 | 6 | 9.8 | **<0.001** |
| Previous individual episode of dengue | 32 | 3.7 | 6 | 7.6 | 9 | 14.7 | **0.001** |
| Comorbidities ^§^ | 435 | 49.9 | 34 | 42.5 | 31 | 50.8 | 0.437 |
| Morbid obesity (body mass index ≥ 40 kg/m^2^) | 19 | 2.2 | 0 | 0.0 | 2 | 3.3 | 0.359 |
| Active smoker ^†^ | 146 | 16.8 | 4 | 5.2 | 12 | 19.7 | **0.022** |
| Fever | 374 | 42.9 | 45 | 56.2 | 59 | 96.7 | **<0.001** |
| Duration of fever (days), µ ± SD | 3.34 | 3.15 | 3.43 | 3.35 | 3.03 | 2.88 | 0.799 |
| Cough | 435 | 49.9 | 36 | 45.0 | 17 | 27.9 | **0.003** |
| Duration of cough (days), µ ± SD | 5.44 | 5.71 | 2.14 | 12.84 | 5.79 | 7.98 | 0.099 |
| Dyspnea/Shortness of breath | 204 | 23.4 | 13 | 16.3 | 13 | 21.3 | 0.332 |
| Duration of dyspnea (days), µ ± SD | 4.14 | 4.50 | 5.44 | 8.43 | 7.75 | 5.25 | 0.376 |
| Body ache ^‡^ | 339 | 38.9 | 32 | 40.0 | 52 | 85.2 | **<0.001** |
| Duration of pain (days), µ ± SD | 3.89 | 3.69 | 4.34 | 5.49 | 2.90 | 2.72 | 0.088 |
| Diarrhea | 179 | 20.2 | 19 | 23.7 | 13 | 21.3 | 0.746 |
| Duration of liquid stools (days), µ ± SD | 2.70 | 2.62 | 4.50 | 3.79 | 2.25 | 3.14 | 0.099 |
| Gut symptoms ^¶^ | 44 | 5.0 | 4 | 5.0 | 13 | 21.3 | **<0.001** |
| Ageusia | 84 | 9.6 | 25 | 31.2 | 11 | 18.0 | **<0.001** |
| Duration of ageusia (days), µ ± SD | 3.93 | 4.16 | 4.73 | 3.32 | 3.25 | 2.01 | 0.263 |
| Metallic taste (dysgeusia) | 4 | 0.5 | 0 | 0.0 | 2 | 3.3 | 0.068 |
| Anosmia | 67 | 7.7 | 28 | 35.0 | 3 | 4.9 | **<0.001** |
| Duration of anosmia (days), µ ± SD | 4.35 | 4.45 | 4.22 | 3.59 | 1.00 | 1.00 | 0.199 |
| Fatigue | 370 | 42.4 | 38 | 47.5 | 49 | 80.3 | **<0.001** |
| Duration of fatigue (days), µ ± SD | 4.29 | 4.20 | 6.48 | 5.75 | 3.44 | 3.00 | **0.027** |
| Headache | 410 | 47.1 | 31 | 38.7 | 56 | 91.8 | **<0.001** |
| Duration of headache (days), µ ± SD | 3.95 | 3.94 | 4.69 | 5.61 | 3.02 | 2.74 | 0.324 |
| Retro-orbital pain | 26 | 3.0 | 1 | 1.2 | 17 | 27.9 | **<0.001** |
| URTI symptoms ^#^ | 459 | 52.6 | 31 | 38.7 | 20 | 32.8 | **0.001** |
| Duration of rhinorrhea (days), µ ± SD | 4.45 | 4.47 | 5.33 | 3.69 | 2.10 | 0.91 | **0.036** |
| Duration of sore throat (days), µ ± SD | 4.17 | 3.98 | 4.00 | 3.27 | 6.20 | 8.22 | 0.995 |
| Presentation > 3 days after symptom onset | 481 | 57.2 | 54 | 70.1 | 24 | 40.0 | **0.002** |
| Time elapsed since symptom onset (days), µ ± SD | 6.27 | 6.25 | 7.54 | 6.50 | 4.18 | 4.57 | **<0.001** |
| Need for physical examination at presentation | 131 | 15.0 | 8 | 10.1 | 19 | 31.1 | **0.001** |
| Dry cough at presentation | 10 | 1.1 | 2 | 2.6 | 1 | 1.6 | 0.315 |
| Anxiety at presentation | 17 | 1.9 | 0 | 0.0 | 0 | 0.0 | 0.459 |
| Frontal temperature (°C), µ ± SD | 37.11 | 0.92 | 36.98 | 0.99 | 37.33 | 1.27 | 0.3375 |
| Cardiac rate (pulses per minute), µ ± SD | 86.84 | 16.46 | 86.38 | 16.80 | 89.89 | 18.60 | 0.520 |
| Respiratory rate (cycles per minute), µ ± SD | 17.56 | 4.88 | 17.39 | 5.69 | 18.09 | 4.98 | 0.479 |
| SpO_2_ (%), µ ± SD | 97.85 | 1.08 | 97.23 | 1.47 | 97.72 | 1.11 | **0.002** |
| Hospitalization | 13 | 1.5 | 14 | 17.5 | 5 | 8.2 | **<0.001** |
| Length of Stay (days), µ ± SD | 1.4 | 0.7 | 9.9 | 7.1 | 1.0 | 0.7 | **<0.001** |
| Data are numbers, column percentages, and *P*-values for Chi2 or Fisher’s exact tests, unless specified as means, standard deviations, and *P*-values for Kruskal-Wallis tests. ^$^15: Urgent Medical Aid Service (SAMU); ^§^diabetes, hypertension, cardiovascular disease, chronic obstructive pulmonary disease, asthma, or cancer; †Current smoker, as compared to never smoker and past smoker; ^‡^muscle pain or backache with tightness and/or stiffness; ^¶^nausea, vomiting, dyspepsia, eructation or abdominal pain ^#^sore throat, runny nose, nasal congestion, or sneezing. | | | | | | | |

**S3 Table. Bivariable multinomial logistic regression distinguishing the crude predictors of COVID-19 and dengue from those of other febrile illnesses among 1,013 participants who consulted a COVID-19 screening center in Saint-Pierre (Reunion Island) during the COVID-19 dengue co-epidemic from 23 March to 10 May 2020**

| Outcomes (vs other febrile illnesses as controls*) | COVID-19 (n = 80) | | | | | Dengue (n = 61) | | | | |
| --- | --- | --- | --- | --- | --- | --- | --- | --- | --- | --- |
| Predictors | **n** | **CIR, (%)** | **cOR** | **95% CI** | ***P* value** | **n** | **CIR, (%)** | **cOR** | **95% CI** | ***P* value** |
| Gender |  |  |  |  |  |  |  |  |  |  |
| Male | 33 | 8.11 | 1 |  |  | 31 | 7.62 | 1 |  |  |
| Female | 47 | 7.76 | 0.92 | 0.57 to 1.47 | 0.738 | 30 | 4.95 | 0.63 | 0.37 to 1.06 | 0.079 |
| Age, years |  |  |  |  |  |  |  |  |  |  |
| 0 to 30 (Q1) | 28 | 9.69 | 1.11 | 0.57 to 2.15 | 0.748 | 8 | 2.77 | **0.34** | **0.13 to 0.83** | **0.018** |
| 31 to 41 (Q2) | 10 | 3.53 | **0.41** | **0.17 to 0.93** | **0.033** | 26 | 9.19 | 1.13 | 0.57 to 2.24 | 0.715 |
| 42 to 54 (Q3) | 27 | 10.34 | 1.23 | 0.63 to 2.39 | 0.542 | 13 | 4.98 | 0.63 | 0.29 to 1.39 | 0.255 |
| 55 to 94 (Q4) | 15 | 8.33 | 1 |  |  | 14 | 7.78 | 1 |  |  |
| Health care worker |  |  |  |  |  |  |  |  |  |  |
| No | 66 | 9.44 | 1 |  |  | 47 | 6.72 | 1 |  |  |
| Yes | 14 | 4.46 | **0.43** | **0.24 to 0.79** | **0.006** | 14 | 4.46 | 0.61 | 0.33 to 1.13 | 0.115 |
| Patient advised by attending physician or 15 |  |  |  |  |  |  |  |  |  |  |
| No | 22 | 5.46 | 1 |  |  | 19 | 4.71 | 1 |  |  |
| Yes | 58 | 9.51 | 1.30 | 0.95 to 1.77 | 0.097 | 42 | 6.89 | **1.84** | **1.19 to 2.86** | **0.006** |
| Contact with a COVID-19 positive case |  |  |  |  |  |  |  |  |  |  |
| No | 38 | 5.18 | 1 |  |  | 55 | 7.49 | 1 |  |  |
| Yes | 42 | 15.05 | **3.07** | **1.22 to 4.88** | **< 0.001** | 6 | 2.15 | **0.30** | **0.12 to 0.71** | **0.006** |
| Return from travel abroad < 15 days |  |  |  |  |  |  |  |  |  |  |
| No | 37 | 4.87 | 1 |  |  | 55 | 7.24 | 1 |  |  |
| Yes | 42 | 16.80 | **3.75** | **2.34 to 6.00** | **< 0.001** | 6 | 2.40 | **0.36** | **0.15 to 0.85** | **0.020** |
| Previous individual episode of dengue |  |  |  |  |  |  |  |  |  |  |
| No | 73 | 7.56 | 1 |  |  | 52 | 5.39 | 1 |  |  |
| Yes | 6 | 12.77 | 2.16 | 0.87 to 5.33 | 0.096 | 9 | 19.15 | **4.54** | **2.05 to 10.02** | **< 0.001** |
| Active smoker ^†^ |  |  |  |  |  |  |  |  |  |  |
| No | 73 | 8.63 | 1 |  |  | 49 | 5.79 | 1 |  |  |
| Yes | 4 | 2.47 | **0.27** | **0.09 to 0.76** | **0.015** | 12 | 7.41 | 1.21 | 0.63 to 2.34 | 0.562 |
| Fever |  |  |  |  |  |  |  |  |  |  |
| No | 3 | 6.55 | 1 |  |  | 2 | 0.37 | 1 |  |  |
| Yes | 45 | 9.41 | **1.71** | **1.07 to 2.71** | **0.023** | 59 | 12.34 | **39.20** | **9.51 to 161.57** | **< 0.001** |
| Cough |  |  |  |  |  |  |  |  |  |  |
| No | 44 | 8.38 | 1 |  |  | 44 | 8.38 | 1 |  |  |
| Yes | 36 | 7.38 | 1 | 0.51 to 1.30 | 0.404 | 17 | 3.48 | **0.39** | **0.21 to 0.69** | **0.001** |
| Dyspnea/Shortness of breath |  |  |  |  |  |  |  |  |  |  |
| No | 67 | 8.56 | 1 |  |  | 48 | 6.13 | 1 |  |  |
| Yes | 13 | 5.65 | 0.64 | 0.34 to 1.17 | 0.148 | 13 | 5.65 | 0.89 | 0.47 to 1.67 | 0.710 |
| To be continued… |  |  |  |  |  |  |  |  |  |  |
| Body ache ^‡^ |  |  |  |  |  | 9 | 1.53 |  |  |  |
| No | 48 | 8.14 | 1 |  |  | 52 | 12.29 | 1 |  |  |
| Yes | 32 | 7.57 | 1.05 | 0.65 to 1.67 | 0.844 |  |  | **9.08** | **4.41 to 18.68** | **< 0.001** |
| Diarrhea |  |  |  |  |  | 48 | 5.97 |  |  |  |
| No | 61 | 7.59 | 1 |  |  | 13 | 6.25 | 1 |  |  |
| Yes | 19 | 9.13 | 1.23 | 0.71 to 2.11 | 0.453 |  |  | 1.07 | 0.56 to 2.02 | 0.836 |
| Gut symptoms ^¶^ |  |  |  |  |  |  |  |  |  |  |
| No | 76 | 7.98 | 1 |  |  | 48 | 5.04 | 1 |  |  |
| Yes | 4 | 6.56 | 0.99 | 0.34 to 2.83 | 0.986 | 13 | 21.31 | **5.10** | **2.57 to 10.10** | **< 0.001** |
| Ageusia |  |  |  |  |  |  |  |  |  |  |
| No | 55 | 6.16 | 1 |  |  | 50 | 5.60 | 1 |  |  |
| Yes | 25 | 20.83 | **4.26** | **2.52 to 7.19** | **< 0.001** | 11 | 9.17 | **2.06** | **1.03 to 4.12** | **0.040** |
| Metallic taste (dysgeusia) |  |  |  |  |  |  |  |  |  |  |
| No | 80 | 7.94 | 1 |  |  | 59 | 5.86 | 1 |  |  |
| Yes | 0 | 0.00 | N.A |  | 0.544 | 2 | 33.3 | 7.36 | 1.32 to 40.99 | 0.008 |
| Anosmia |  |  |  |  |  |  |  |  |  |  |
| No | 52 | 5.68 | 1 |  |  | 58 | 6.34 | 1 |  |  |
| Yes | 28 | 28.57 | **6.47** | **3.83 to 10.91** | **< 0.001** | 3 | 3.06 | 0.62 | 0.18 to 2.04 | 0.432 |
| Fatigue |  |  |  |  |  |  |  |  |  |  |
| No | 42 | 7.55 | 1 |  |  | 12 | 2.16 | 1 |  |  |
| Yes | 38 | 8.32 | 1.23 | 0.77 to 1.94 | 0.381 | 49 | 10.72 | **5.54** | **2.90 to 10.57** | **< 0.001** |
| Headache |  |  |  |  |  |  |  |  |  |  |
| No | 49 | 9.51 | 1 |  |  | 5 | 0.97 | 1 |  |  |
| Yes | 31 | 6.24 | 0.71 | 0.44 to 1.14 | 0.155 | 56 | 11.27 | **12.59** | **4.99 to 31.75** | **< 0.001** |
| Retro-orbital pain |  |  |  |  |  |  |  |  |  |  |
| No | 79 | 8.15 | 1 |  |  | 44 | 4.54 | 1 |  |  |
| Yes | 1 | 2.27 | 0.41 | 0.05 to 3.08 | 0.401 | 17 | 38.64 | **12.57** | **6.35 to 24.88** | **< 0.001** |
| URTI symptoms ^#^ |  |  |  |  |  |  |  |  |  |  |
| No | 49 | 9.74 | 1 |  |  | 41 | 8.15 | 1 |  |  |
| Yes | 31 | 6.08 | **0.57** | **0.35 to 0.91** | **0.019** | 20 | 3.92 | **0.44** | **0.25 to 0.76** | **0.003** |
| Presentation > 3 days after symptom onset |  |  |  |  |  |  |  |  |  |  |
| No | 23 | 5.49 | 1 |  |  | 36 | 8.59 | 1 |  |  |
| Yes | 54 | 9.66 | **1.76** | **1.05 to 2.92** | **0.029** | 24 | 4.29 | **0.50** | **0.29 to 0.85** | **0.011** |
| *Other non-COVID-19 non dengue febrile illnesses. Data are numbers, cumulative incidence rates (CIR) expressed as percentages, crude odd ratios (cOR), 95% confidence intervals (95% CI) and *P*-values for Wald tests. ^$^15: Urgent Medical Aid Service (SAMU); †Current smokers, as compared to never smokers and past smokers; ^‡^muscle pain or backache with tightness and/or stiffness; ^¶^nausea, vomiting, dyspepsia, eructation or abdominal pain; ^#^sore throat, runny nose, nasal congestion, or sneezing. N.A: not assessed (incalculable). | | | | | | | | | | |

| S4 Table. Full multinomial logistic regression model distinguishing independent predictors of COVID-19 and dengue from those of other febrile illnesses among 967 participants who consulted a screening center in Saint-Pierre (Reunion Island) during the COVID-19 dengue co-epidemics from 23 March to 10 May 2020 | | | | | | |
| --- | --- | --- | --- | --- | --- | --- |
| Outcomes (vs other febrile illnesses as controls*) | **COVID-19 (n = 80)** | | | **Dengue (n = 61)** | | |
| Predictors | **aOR** | **95% CI** | ***P* value** | **aOR** | **95% CI** | ***P* value** |
| Female Gender | 0.92 | 0.54 to 1.55 | 0.754 | 0.87 | 0.42 to 1.76 | 0.693 |
| Age, years |  |  |  |  |  |  |
| 0 to 30 (Q1) | 0.94 | 0.42 to 2.11 | 0.883 | 0.29 | 0.08 to 0.93 | 0.037 |
| 31 to 41 (Q2) | 0.62 | 0.25 to 1.51 | 0.291 | 1.51 | 0.56 to 3.99 | 0.408 |
| 42 to 54 (Q3) | 1.66 | 0.77 to 3.57 | 0.196 | 0.66 | 0.22 to 1.98 | 0.463 |
| 55 to 94 (Q4) | 1 |  |  | 1 |  |  |
| Health care worker | 0.54 | 0.20 to 1.47 | 0.229 | 1.13 | 0.38 to 3.37 | 0.824 |
| Patient advised by attending physician or 15 | 1.07 | 0.51 to 2.24 | 0.851 | 1.24 | 0.64 to 2.40 | 0.518 |
| Contact with a COVID-19 positive case | 5.28 | **2.80 to 9.93** | **< 0.001** | 1.67 | 0.50 to 5.56 | 0.402 |
| Return from travel abroad < 15 days | 3.42 | **1.93 to 6.04** | **< 0.001** | 0.43 | 0.14 to 1.28 | 0.129 |
| Previous individual episode of dengue | 2.47 | 0.55 to 10.95 | 0.234 | 4.15 | **1.15 to 14.86** | **0.029** |
| Active smoker ^†^ | 0.31 | **0.11 to 0.86** | **0.024** | 1.25 | 0.50 to 3.14 | 0.630 |
| Fever | 2.09 | **1.01 to 4.28** | **0.045** | 17.62 | **3.90 to 79.54** | **< 0.001** |
| Cough | 0.85 | 0.47 to 1.52 | 0.586 | 0.32 | **0.15 to 0.65** | **0.002** |
| Body ache ^‡^ | 0.80 | 0.40 to 1.58 | 0.518 | 2.71 | **1.02 to 7.15** | **0.044** |
| Diarrhea | 0.95 | 0.46 to 1.94 | 0.891 | 0.53 | 0.22 to 1.26 | 0.149 |
| Gut symptoms ^¶^ | 1.72 | 0.54 to 5.35 | 0.353 | 3.26 | **1.17 to 9.03** | **0.023** |
| Ageusia | 1.07 | 0.43 to 2.61 | 0.889 | 2.17 | 0.71 to 6.55 | 0.170 |
| Anosmia | 6.85 | **2.73 to 17.16** | **< 0.001** | 0.26 | 0.04 to 1.60 | 0.146 |
| Fatigue | 1.13 | 0.55 to 2.29 | 0.737 | 1.82 | 0.75 to 4.39 | 0.182 |
| Headache | 0.87 | 0.49 to 1.51 | 0.616 | 3.18 | **1.04 to 9.74** | **0.042** |
| Retro-orbital pain | 0.40 | 0.04 to 3.64 | 0.417 | 5.54 | **2.42 to 14.07** | **< 0.001** |
| URTI symptoms ^#^ | 0.57 | **0.33 to 0.99** | **0.045** | 0.56 | 0.27 to 1.16 | 0.120 |
| Presentation > 3 days after symptom onset | 1.69 | 0.92 to 3.08 | 0.087 | 0.60 | 0.31 to 1.14 | 0.120 |
| *Other non-COVID-19 non dengue febrile illnesses. Data are numbers, cumulative incidence rates (CIR) expressed as percentages, adjusted odd ratios (aOR), 95% confidence intervals (95% CI) and *P*-values for Wald tests. ^$^15: Urgent Medical Aid Service (SAMU); †Current smokers, as compared to never smokers and past smokers ^‡^muscle pain or backache with tightness and/or stiffness; ^¶^nausea, vomiting, dyspepsia, eructation or abdominal pain; ^#^sore throat, runny nose, nasal congestion, or sneezing. 46 Participants with missing data excluded from the model (1,013-967). The indicators of performance of the model are as follows: Bayesian information criterion -5541, Goodness of fit chi-2 test’s probability 0.69, areas under the receiver operating characteristic curves 0.83 (95%CI 0.77 to 0.88) and 0.93 (95%CI 0.90 to 0.96), respectively. | | | | | | |

**S5 Table. Final weighted parsimonious multinomial logistic regression model distinguishing independent predictors of COVID-19 and dengue from those of other febrile illnesses among 969 participants who consulted at a COVID-19 screening center in Saint-Pierre (Reunion Island) during the COVID-19 dengue co-epidemics from 23 March to 10 May 2020**

| Outcomes (versus other febrile illnesses as controls*) | COVID-19 (n = 73) | | | | Dengue (n = 60) | | | |
| --- | --- | --- | --- | --- | --- | --- | --- | --- |
| Predictors | **CIR, %** | **aOR** | **95% CI** | ***P* values** | **CIR, %** | **aOR** | **95% CI** | ***P* values** |
| Contact with a COVID-19 positive case | 18.58 | **10.31** | **4.38 to 24.25** | **< 0.001** | 7.43 | 3.31 | 0.35 to 27.10 | 0.304 |
| Return from travel abroad < 15 days | 18.96 | **8.88** | **4.21 to 18.70** | **< 0.001** | 7.93 | 1.41 | 0.24 to 7.99 | 0.699 |
| Previous individual episode of dengue | 4.10 | 3.94 | 0.75 to 20.65 | 0.104 | 31.67 | **11.48** | **1.54 to 85.45** | **0.017** |
| Active smoker † | 1.61 | **0.32** | **0.10 to 1.00** | **0.051** | 20.92 | 1.92 | 0.19 to 18.81 | 0.573 |
| Cough | 4.15 | 0.77 | 0.32 to 1.82 | 0.545 | 6.30 | 0.58 | 0.13 to 2.49 | 0.460 |
| Body ache ^‡^ | 4.60 | 1.32 | 0.57 to 3.02 | 0.514 | 16.79 | 6.20 | 0.81 to 47.27 | 0.078 |
| Anosmia | 9.75 | **5.16** | **1.94 to 13.69** | **0.001** | 10.1 | 1.21 | 0.14 to 10.47 | 0.862 |
| Headache | 3.78 | 1.11 | 0.46 to 2.66 | 0.809 | 16.80 | **36.06** | **9.83 to 132.23** | **< 0.001** |
| Retro-orbital pain | 1.05 | 0.35 | 0.02 to 5.49 | 0.453 | 26.91 | 0.99 | 0.15 to 6.52 | 0.996 |
| URTI symptoms ^#^ | 3.61 | 0.56 | 0.24 to 1.32 | 0.185 | 6.82 | **0.35** | **0.06 to 1.90** | 0.224 |
| Presentation > 3 days after symptom onset | 6.25 | 1.65 | 0.72 to 3.75 | 0.234 | 8.28 | 0.87 | 0.11 to 6.42 | 0.891 |
| Multinomial logistic regression model with other non COVID-19 non dengue febrile illnesses*, taken as controls. Data are numbers, cumulative incidence rates (CIR) expressed as percentages, adjusted odd ratios (aOR), 95% confidence intervals (95% CI) and *P*-values for Wald tests. †Current smoker, as compared to never smoker and past smoker; ‡ muscle pain or backache with tightness and/or stiffness; ^#^sore throat, runny nose, nasal congestion, or sneezing. Post-estimation metrics are not possible after *svyset* for multinomial logistic regression. In total, 44 Participants with missing data excluded from the model (1,013 – 969). | | | | | | | | |

| **S6 Table. Benchmarking of goodness-of-fit and discrimination metrics for five multinomial logistic regression models distinguishing COVID-19 and dengue from other febrile illnesses among the participants who consulted at a screening center in Saint-Pierre (Reunion Island) during the COVID-19 dengue co-epidemic from 23 March to 10 May 2020** | | | | | | | | | | | |
| --- | --- | --- | --- | --- | --- | --- | --- | --- | --- | --- | --- |
| **Multinomial logistic regression model** | **Full main-effect**  **20-covariate model^1^** | | **13-covariate model**  **(+ age and gender)^2^** | | | **12-covariate model**  **(+ age)^3^** | | **11-covariate**  **final model^4^** | | **Minimal-effect**  **9-covariate model^5^** | |
| **Criteria of validity** | | | | | | | | | | | |
| **Representativeness, n (%)** | 967 (95.4) | 5 | 969 (95.6) | | 2 | 969 (95.6) | 2 | **969 (95.6)** | **2** | 972 (95.9) | 1 |
| **Mac Fadden pseudo R2** | 0.3405 | 1 | 0.2864 | | 2 | 0.2859 | 3 | **0.2630** | **4** | 0.2250 | 5 |
| **Aïkake information criterion** | 0.796 | 1 | 0.805 | | 4 | 0.799 | 2 | **0.803** | **3** | 0.830 | 5 |
| **Bayesian information criterion** | - 5541 | 5 | - 5649 | | 4 | - 5669 | 3 | - **5709** | **2** | - 5733 | 1 |
| **Deviance** | 631 | 1 | 683 | | 4 | 684 | 3 | **706** | **2** | 746 | 5 |
| **Hosmer-Lemeshow goodness-of-fit test** | 0.689 | 4 | 0.694 | | 3 | 0.711 | 2 | **0.313** | **5** | 0.823 | 1 |
| **COVID-19 bootstrap AUC estimate** | † 0.8296 | 1 | 0.8188 | | 3 | † 0.8197 | 2 | ***0.8008** | **4** | **0.7719 | 5 |
| **COVID-19 bootstrap AUC standard error** | 0.0260 | 1 | 0.0263 | | 2 | 0.0264 | 3 | **0.0297** | **4** | 0.0298 | 5 |
| **Dengue bootstrap AUC estimate** | ‡ 0.9341 | 1 | **0.8959 | | 2 | **0.8955 | 3 | ****0.8828** | **4** | **0.8721 | 5 |
| **Dengue bootstrap AUC standard error** | 0.0148 | 1 | 0.0194 | | 4 | 0.0192 | 3 | **0.0191** | **2** | 0.0203 | 5 |
| **Average ranking (1 to 5)^a^** |  | 2.1 |  | | 3.0 |  | 2.6 |  | **3.2** |  | 3.8 |
| Pr. (minimizing information loss, AIC) **^b^** | 1.0000 | 1 | 0.99551 | | 4 | 0.9985 | 2 | **0.9965** | **3** | 0.9091 | 5 |
| Pr. (fulfilling principle of parsimony, BIC)^c^ | 0.6003 | 5 | 0.8305 | | 4 | 0.8983 | 3 | **0.9979** | **1** | 0.9625 | 2 |
| **Total score a+b+c (model ranking)** | 8.1 (3) | | | 11.0 (5) | | 7.6 (2) | | **7.2 (1)** | | 10.8 (4) | |
| Model AUCs were compared two-by-two using DeLong method taking Model 1(*) or Model 4 (†) as reference model. **‡ *P-*value< 0.01. *† 0.01<P-value<0.05. The interpretation of the discriminative ability can be evaluated using both De Long method and subtracting its standard error to the AUC. The 20-item model was superior to all other models for dengue and to the 11-item model for COVID-19. Adding age to the 11-item COVID-19 model improved discrimination without changing interpretation. | | | | | | | | | | | |

| **S7 Table. Calibration of the COVID score across the deciles of predicted risk by across the deciles of predicted risk by the Hosmer-Lemeshow goodness-of-fit chi square test among 969 participants who consulted at a screening center in Saint-Pierre (Reunion Island) during the COVID-19 dengue co-epidemics from 23 March to 10 May 2020** | | | | | | | | | |
| --- | --- | --- | --- | --- | --- | --- | --- | --- | --- |
| **Decile** | **Predicted risk** | **Observed_0** | **Predicted_0** | **Observed_1** | **Predicted_1** | **Total** | **Ratio Obs. to Pre.** | **Chi2 value (ddl)** | **Pr.(chi2)** |
| **1** | 0.0171 | 182 | 181.6 | 2 | 2.4 | 184 | 0.8333 | 0.07 (1) | **0.791** |
| **4** | 0.0242 | 225 | 227.4 | 8 | 5.6 | 233 | 1.4286 | 1.05 (1) | **0.305** |
| **5** | 0.0341 | 114 | 114.0 | 4 | 4.0 | 118 | 1.0000 | 0.00 (1) | **1.000** |
| **6** | 0.0478 | 50 | 49.5 | 2 | 2.5 | 52 | 0.8000 | 0.11 (1) | **0.740** |
| **7** | 0.0667 | 159 | 159.6 | 12 | 11.4 | 171 | 1.0526 | 0.03 (1) | **0.862** |
| **8** | 0.0924 | 70 | 70.8 | 8 | 7.2 | 78 | 1.1111 | 0.10 (1) | **0.752** |
| **9** | 0.1710 | 41 | 39.6 | 6 | 7.4 | 47 | 0.8108 | 0.31 (1) | **0.578** |
| **10** | 0.7770 | 55 | 53.6 | 31 | 32.4 | 86 | 0.9568 | 0.10 (1) | **0.752** |
| **Total** | 1.0000 | 896 | 896 | 73 | 73 | 969 | 1.0000 | 1.75 (6) | **0.942** |
| Calibration (the adequacy between predicted and observed events) was assessed using the Hosmer-Lemeshow goodness-of-fit chi square test overall and individually across the deciles of a predicted risk. Calibration was deemed satisfactory for 0.05<*P*-value<0.5 and excellent for *P*-value>0.5. Each test was conservative and rejected the alternative hypothesis of independency between the predicted risk and the COVID-19 event (*P*>0.05). | | | | | | | | | |

| **S8 Table. Calibration of the dengue score across the deciles of predicted risk by the Hosmer-Lemeshow Goodness-of-fit Chi-square test among 969 participants who consulted at a screening center in Saint-Pierre (Reunion Island) during the COVID-19 dengue co-epidemics from 23 March to 10 May 2020** | | | | | | | | | |
| --- | --- | --- | --- | --- | --- | --- | --- | --- | --- |
| **Decile** | **Predicted risk** | **Observed_0** | **Predicted_0** | **Observed_1** | **Predicted_1** | **Total** | **Ratio Obs. to Pre.** | **Chi2 value (ddl)** | **Pr.(chi2)** |
| **1** | 0.0018 | 99 | 98.8 | 0 | 0.2 | 99 | 0.0000 | 0.20 (1) | **0.655** |
| **2** | 0.0039 | 164 | 163.5 | 0 | 0.5 | 164 | 0.0000 | 0.50 (1) | **0.479** |
| **3** | 0.0058 | 89 | 88.5 | 0 | 0.5 | 89 | 0.0000 | 0.50 (1) | **0.479** |
| **4** | 0.0084 | 36 | 35.7 | 0 | 0.3 | 36 | 0.0000 | 0.30 (1) | **0.584** |
| **5** | 0.0178 | 140 | 141.9 | 4 | 2.1 | 144 | 1.9048 | 1.74 (1) | **0.187** |
| **6** | 0.0259 | 66 | 67.2 | 3 | 1.8 | 69 | 1.6667 | 0.82 (1) | **0.365** |
| **7** | 0.0537 | 86 | 86.2 | 4 | 3.8 | 90 | 1.0536 | 0.01 (1) | **0.920** |
| **8** | 0.1084 | 126 | 121.2 | 7 | 11.8 | 133 | 0.5932 | 2.14 (1) | **0.143** |
| **9** | 0.1509 | 46 | 47.5 | 10 | 8.5 | 56 | 1.7614 | 0.31 (1) | **0.578** |
| **10** | 0.8450 | 57 | 58.5 | 12 | 30.5 | 89 | 0.3934 | 0.11 (1) | **0.740** |
| **Total** | 1.0000 | 909 | 909 | 60 | 60 | 969 | 1.0000 | 6.85 (8) | **0.553** |
| Calibration (the adequacy between predicted and observed events) was assessed using the Hosmer-Lemeshow Goodness-of-fit Chi-square test overall and individually across the deciles of predicted risk. Calibration was deemed satisfactory for 0.05<*P*-value<0.5 and excellent for *P* values>0.5. Each test was conservative and rejected the alternative hypothesis of independency between the predicted risk and the COVID-19 event (*P*>0.05). | | | | | | | | | |

| **S9 Table. Benchmarking of goodness-of-fit metrics for three models and two scores distinguishing COVID-19 and dengue predictors from those of other febrile illnesses among 969 participants who consulted at a screening center in Saint Pierre (Reunion Island) during the COVID-19 dengue co-epidemic from 23 March to 10 May 2020** | | | | | | | | | | |
| --- | --- | --- | --- | --- | --- | --- | --- | --- | --- | --- |
| **Multinomial logistic regression model** | **11-covariate**  **final model^4^** | | **11-covariate**  **COVID model^6^** | | **11-covariate**  **COVID score^7^** | | **11-covariate**  **dengue model^8^** | | **11-covariate**  **dengue score^9^** | |
| **Criteria of validity^5^** | | |  | | | |  | | | |
| **Mac Fadden pseudo R2** | 0.2630 | 3 | 0.1413 | 5 | **0.2092** | **4** | 0.3113 | 1 | **0.2936** | **2** |
| **Aïkake information criterion** | 0.803 | 5 | 0.712 | 4 | **0.427** | **3** | 0.345 | 2 | **0.332** | **1** |
| **Bayesian information criterion** | - 5709 | 5 | - 5914 | 4 | - **6239** | **3** | - 6270 | 2 | - **6331** | **1** |
| **Deviance residuals** | 706 | 5 | 665 | 4 | **409** | **3** | 309 | 1 | **318** | **2** |
| **Hosmer-Lemeshow goodness-of-fit test** | 0.313 | 4 | 0.007 | 5 | **0.942** | **1** | 0.876 | 2 | **0.560** | **3** |
| **Bootstrap AUC estimate** | 0.9880 | 1 | 0.7132 | 5 | **0.7510** | **4** | 0.8844 | 2 | **0.8607** | **3** |
| **Bootstrap AUC standard error** | 0.0004 | 1 | 0.0357 | 5 | **0.0339** | **4** | 0.0196 | 2 | **0.0211** | **3** |
| **Average ranking (1 to 5)** |  | 3.4 |  | 4.6 |  | **3.1** |  | 1.7 |  | **2.1** |
| **Ranking** |  | 4 |  | 5 |  | **3** |  | 1 |  | **2** |
| AUCs of models and scores were compared two-by-two using DeLong method taking model 6(*) or model 8(†) as reference model. **‡ P< 0.01. *† 0.01<P<0.05. The interpretation of the discriminative ability can be evaluated using both De Long method and subtracting its standard error to the AUC. There was no significant difference in discrimination performance between each score and each of the reference models. | | | | | | | | | | |

| **S10 Table. Diagnostic performance of the COVID score to predict COVID-19 among 969 participants who consulted at a screening center in Saint Pierre (Reunion Island) during the COVID-19 dengue co-epidemics from 23 March to 10 May 2020** | | | | | | | | | | | | | | |
| --- | --- | --- | --- | --- | --- | --- | --- | --- | --- | --- | --- | --- | --- | --- |
| **Score value** | **Sen** | | **95% CI** | **Spe** | **95% CI** | **LR+** | **95% CI** | **LR-** | **95% CI** | **PPV** | **95% CI** | **NPV** | **95% CI** | **Accuracy** |
| -5 | 1.00 | 0.95 to 1.00 | | 0.00 | 0.00 to 0.41 | 0.99 | 0.98 to 1.01 | 12.10 | 0.24 to 607 | 0.07 | 0.06 to 0.07 | - | 0.02 to 1.00 | 0.00 |
| -4 | 1.00 | 0.95 to 1.00 | | 0.00 | 0.00 to 0.41 | 1.00 | 0.97 to 1.01 | 12.10 | 0.24 to 607 | 0.07 | 0.06 to 0.07 | - | 0.22 to 1.00 | 0.00 |
| -3 | 1.00 | 0.95 to 1.00 | | 0.03 | 0.02 to 0.04 | 1.02 | 1.00 to 1.04 | 0.24 | 0.01 to 3.86 | 0.07 | 0.07 to 0.07 | 1.00 | 0.78 to 1.00 | 0.03 |
| **-2** | 1.00 | 0.95 to 1.00 | | 0.07 | 0.06 to 0.09 | 1.07 | 1.04 to 1.10 | **0.09** | **0.01 to 1.50** | 0.07 | 0.07 to 0.07 | 1.00 | 0.90 to 1.00 | 0.07 |
| -1 | 0.99 | 0.93 to 1.00 | | 0.09 | 0.07 to 0.11 | 1.09 | 1.05 to 1.12 | **0.15** | **0.02 to 1.05** | 0.07 | 0.07 to 0.08 | 0.99 | 0.93 to 1.00 | 0.09 |
| **0** | 0.97 | 0.90 to 1.00 | | 0.20 | 0.18 to 0.23 | 1.22 | 1.16 to 1.28 | 0.13 | 0.13 to 0.53 | 0.08 | 0.08 to 0.09 | 0.99 | 0.96 to 1.00 | 0.19 |
| 1 | 0.86 | 0.76 to 0.93 | | 0.45 | 0.42 to 0.49 | 1.58 | 1.42 to 1.76 | 0.30 | 0.17 to 0.54 | 0.10 | 0.09 to 0.11 | 0.98 | 0.96 to 0.99 | 0.43 |
| 2 | 0.81 | 0.70 to 0.89 | | 0.58 | 0.55 to 0.61 | 1.93 | 1.69 to 2.21 | 0.33 | 0.21 to 0.53 | 0.12 | 0.11 to 0.14 | 0.98 | 0.96 to 0.99 | 0.55 |
| **3** | **0.78** | **0.67 to 0.87** | | **0.64** | **0.61 to 0.67** | **2.15** | **1.85 to 2.50** | **0.34** | **0.22 to 0.53** | **0.14** | **0.12 to 0.15** | **0.98** | **0.96 to 0.98** | **0.60** |
| 4 | 0.62 | 0.50 to 0.73 | | 0.81 | 0.79 to 0.84 | 3.33 | 2.65 to 4.18 | 0.47 | 0.35 to 0.63 | 0.19 | 0.16 to 0.23 | 0.97 | 0.96 to 0.98 | 0.76 |
| 5 | 0.51 | 0.39 to 0.63 | | 0.89 | 0.87 to 0.91 | 4.73 | 3.52 to 6.35 | 0.55 | 0.44 to 0.70 | 0.26 | 0.20 to 0.32 | 0.96 | 0.95 to 0.97 | 0.84 |
| 6 | 0.49 | 0.37 to 0.61 | | 0.91 | 0.89 to 0.93 | 5.32 | 3.90 to 7.26 | 0.56 | 0.45 to 0.70 | 0.28 | 0.22 to 0.35 | 0.96 | 0.95 to 0.97 | 0.85 |
| 7 | 0.42 | 0.31 to 0.55 | | 0.94 | 0.92 to 0.95 | 6.92 | 4.78 to 10.00 | 0.61 | 0.50 to 0.75 | 0.33 | 0.26 to 0.42 | 0.96 | 0.95 to 0.97 | 0.88 |
| **8** | 0.34 | 0.31 to 0.37 | | 0.97 | 0.96 to 0.98 | **11.36** | **6.97 to 18.50** | 0.68 | 0.57 to 0.80 | 0.45 | 0.34 to 0.57 | 0.95 | 0.95 to 0.96 | 0.91 |
| 9 | 0.34 | 0.24 to 0.46 | | 0.99 | 0.98 to 0.99 | **23.60** | **12.60 to 44.20** | 0.67 | 0.57 to 0.79 | 0.63 | 0.48 to 0.76 | 0.95 | 0.95 to 0.96 | 0.92 |
| **10** | 0.32 | 0.21 to 0.43 | | **0.99** | **0.98 to 0.99** | **25.66** | **21.10 to 43.40** | 0.69 | 0.59 to 0.81 | **0.65** | **0.48 to 0.79** | 0.95 | 0.94 to 0.96 | 0.93 |
| 11 | 0.19 | 0.11 to 0.30 | | 1.00 | 0.99 to 1.00 | **42.96** | **14.50 to 127.00** | 0.81 | 0.73 to 0.91 | 0.76 | 0.51 to 0.90 | 0.94 | 0.94 to 0.95 | 0.93 |
| 13 | 0.10 | 0.04 to 0.19 | | 1.00 | 0.99 to 1.00 | **42.96** | **9.09 to 203.00** | 0.91 | 0.84 to 0.98 | 0.76 | 0.40 to 0.94 | 0.94 | 0.93 to 0.94 | 0.94 |
| 14 | 0.08 | 0.03 to 0.17 | | 1.00 | 1.00 to 1.00 | **158.00** | **8.96 to 2770.00** | 0.91 | 0.85 to 0.98 | 1.00 | 0.39 to 1.00 | 0.94 | 0.93 to 0.94 | 0.94 |
| 15 | 0.00 | 0.00 to 0.05 | | 1.00 | 1.00 to 1.00 | **12.10** | **0.24 to 607.00** | 0.99 | 0.98 to 1.01 | **-** | 0.02 to 0.98 | 0.93 | 0.93 to 0.93 | 0.94 |
| Sen: sensitivity; Spe: specificity; LR+: positive likelihood ratio; LR-: negative likelihood ratio; PPV: positive predictive value; NPV: negative predictive value | | | | | | | | | | | | | | |

| **11 Table. Diagnostic performance of the dengue score to predict dengue among 969 participants who consulted at a screening center in Saint Pierre (Reunion Island) during the COVID-19 dengue co-epidemics from 23 March to 10 May 2020** | | | | | | | | | | | | | |
| --- | --- | --- | --- | --- | --- | --- | --- | --- | --- | --- | --- | --- | --- |
| **Score value** | **Sen** | **95% CI** | **Spe** | **95% CI** | **LR+** | **95% CI** | **LR-** | **95% CI** | **PPV** | **95% CI** | **NPV** | **95% CI** | **Accuracy** |
| -5 | 1.00 | 0.94 to 1.00 | 0.00 | 0.00 to 0.00 | 0.99 | 0.97 to 1.02 | **14.90** | **0.30 to 745.00** | 0.04 | 0.04 to 0.05 | . | 0.02 to 1.00 | 0.00 |
| -4 | 1.00 | 0.94 to 1.00 | 0.00 | 0.00 to 0.01 | 1.00 | 0.97 to 1.02 | **2.98** | **0.15 to 61.50** | 0.04 | 0.04 to 0.05 | 1.00 | 0.26 to 1.00 | 0.00 |
| -3 | 1.00 | 0.94 to 1.00 | 0.03 | 0.02 to 0.04 | 1.02 | 1.00 to 1.05 | **0.28** | **0.02 to 4.56** | 0.05 | 0.04 to 0.05 | 1.00 | 0.82 to 1.00 | 0.03 |
| -2 | 1.00 | 0.94 to 1.00 | 0.11 | 0.09 to 0.13 | 1.11 | 1.08 to 1.15 | **0.08** | **0.00 to 1.19** | 0.05 | 0.05 to 0.05 | 1.00 | 0.95 to 1.00 | 0.11 |
| -1 | 1.00 | 0.94 to 1.00 | 0.20 | 0.17 to 0.22 | 1.23 | 1.19 to 1.28 | **0.04** | **0.00 to 0.66** | 0.05 | 0.05 to 0.06 | 1.00 | 0.97 to 1.00 | 0.19 |
| 0 | 1.00 | 0.94 to 1.00 | 0.29 | 0.26 to 0.32 | 1.40 | 1.33 to 1.46 | **0.03** | **0.00 to 0.45** | 0.06 | 0.06 to 0.06 | 1.00 | 0.98 to 1.00 | 0.28 |
| 1 | 1.00 | 0.94 to 1.00 | 0.39 | 0.36 to 0.42 | 1.62 | 1.53 to 1.71 | **0.02** | **0.00 to 0.34** | 0.07 | 0.07 to 0.07 | 1.00 | 0.99 to 1.00 | 0.37 |
| 2 | 1.00 | 0.94 to 1.00 | 0.43 | 0.39 to 0.46 | 1.73 | 1.63 to 1.84 | **0.02** | **0.00 to 0.30** | 0.08 | 0.07 to 0.08 | 1.00 | 0.99 to 1.00 | 0.41 |
| **3** | **0.97** | **0.88 to 1.00** | **0.52** | 0.49 to 0.56 | 2.03 | 1.87 to 2.21 | **0.06** | **0.02 to 0.25** | 0.09 | 0.08 to 0.09 | **0.99** | **0.98 to 1.00** | 0.50 |
| 4 | 0.93 | 0.83 to 0.98 | 0.58 | 0.55 to 0.61 | 2.23 | 2.01 to 2.47 | 0.11 | 0.04 to 0.30 | 0.09 | 0.09 to 0.10 | 0.99 | 0.98 to 1.00 | 0.56 |
| 5 | 0.88 | 0.77 to 0.95 | 0.65 | 0.62 to 0.68 | 2.55 | 2.24 to 2.90 | 0.18 | 0.09 to 0.36 | 0.11 | 0.09 to 0.12 | 0.99 | 0.98 to 1.00 | 0.63 |
| **6** | **0.83** | **0.72 to 0.92** | **0.72** | **0.69 to 0.75** | **2.97** | **2.53 to 3.44** | **0.23** | **0.13 to 0.41** | **0.12** | **0.11 to 0.14** | **0.99** | **0.98 to 0.99** | **0.69** |
| 7 | 0.82 | 0.70 to 0.91 | 0.75 | 0.72 to 0.78 | 3.24 | 2.75 to 3.82 | 0.25 | 0.14 to 0.42 | 0.13 | 0.11 to 0.15 | 0.99 | 0.98 to 0.99 | 0.72 |
| 8 | 0.73 | 0.60 to 0.84 | 0.83 | 0.81 to 0.86 | 4.39 | 3.55 to 5.41 | 0.32 | 0.21 to 0.49 | 0.17 | 0.14 to 0.20 | 0.99 | 0.98 to 0.99 | 0.80 |
| 9 | 0.70 | 0.56 to 0.81 | 0.89 | 0.86 to 0.91 | 6.18 | 4.83 to 7.90 | 0.34 | 0.23 to 0.50 | 0.22 | 0.18 to 0.27 | 0.98 | 0.98 to 0.99 | 0.85 |
| 10 | 0.53 | 0.40 to 0.66 | 0.94 | 0.92 to 0.95 | 8.51 | 6.02 to 12.00 | 0.50 | 0.38 to 0.65 | 0.28 | 0.22 to 0.36 | 0.98 | 0.97 to 0.98 | 0.90 |
| **11** | 0.33 | 0.22 to 0.47 | **0.98** | **0.96 to 0.99** | **14.43** | **8.29 to 25.10** | 0.68 | 0.57 to 0.82 | 0.40 | 0.28 to 0.54 | 0.97 | 0.96 to 0.97 | 0.94 |
| 12 | 0.33 | 0.22 to 0.47 | 0.98 | 0.97 to 0.99 | **16.83** | **9.42 to 30.10** | 0.68 | 0.65 to 0.71 | 0.44 | 0.31 to 0.58 | 0.97 | 0.96 to 0.97 | 0.94 |
| 13 | 0.30 | 0.19 to 0.43 | 0.99 | 0.98 to 0.99 | **22.72** | **11.50 to 45.00** | 0.71 | 0.68 to 0.74 | **0.51** | **0.35 to 0.68** | 0.97 | 0.96 to 0.97 | 0.94 |
| 14 | 0.17 | 0.08 to 0.29 | 0.99 | 0.98 to 1.00 | **16.83** | **7.11 to 39.90** | 0.84 | 0.75 to 0.94 | 0.44 | 0.25 to 0.65 | 0.96 | 0.96 to 0.97 | 0.95 |
| 15 | 0.13 | 0.06 to 0.25 | 0.99 | 0.99 to 1.00 | **20.20** | **7.24 to 56.30** | 0.87 | 0.79 to 0.96 | 0.49 | 0.25 to 0.73 | 0.96 | 0.96 to 0.96 | 0.95 |
| 16 | 0.03 | 0.00 to 0.12 | 1.00 | 0.99 to 1.00 | **30.30** | **2.79 to 329.00** | 0.97 | 0.92 to 1.01 | 0.59 | 0.12 to 0.94 | 0.96 | 0.96 to 0.96 | 0.96 |
| 18 | 0.02 | 0.00 to 0.09 | 1.00 | 1.00 to 1.00 | **44.80** | **1.84 to 1087.00** | 0.98 | 0.94 to 1.02 | 1.00 | 0.08 to 1.00 | 0.96 | 0.96 to 0.96 | 0.96 |
| Sen: sensitivity; Spe: specificity; LR+: positive likelihood ratio; LR-: negative likelihood ratio; PPV: positive predictive value; NPV: negative predictive value | | | | | | | | | | | | | |

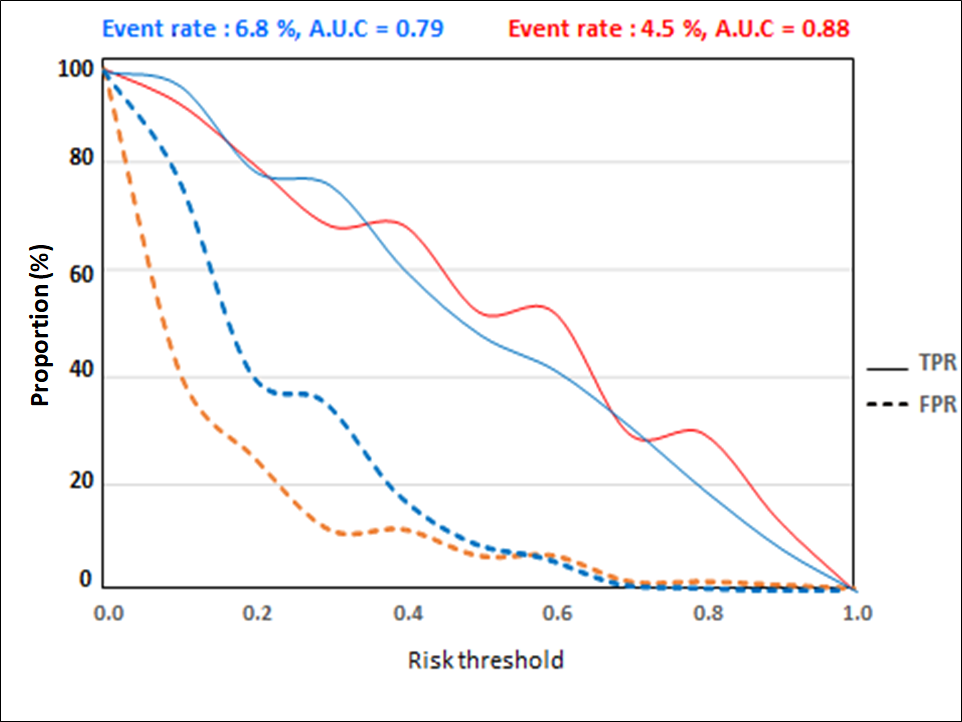

**S1 Figure. Classification plots of the COVIDENGUE scores**

Notes: State-of-art classification plots are given for the COVIDENGUE scores before internal validation by bootstrapping. Event rate = prevalence (%). A.U.C = area under receiver operating characteristic curve; TPR: true positive rate (sensitivity); FPR: false positive rate (100-specifity). TPR and FPR curves (y axis) are given according to risk thresholds, the observed percent of events (x axis). The area under the TPR (sensitivity) curve equals the mean predicted risk of events for events (true positives), whereas the area under the FPR (100-specificity) equals the mean predicted risk of events for non-events (false positives). Overall, the plots mean that the COVID score has a better ability to predict other febrile illnesses, especially at lower risk thresholds (0 to 0.4) and that the dengue score has a better ability to predict infection, especially at higher risk thresholds (0.4 to 1.0) with exception of 0.7).

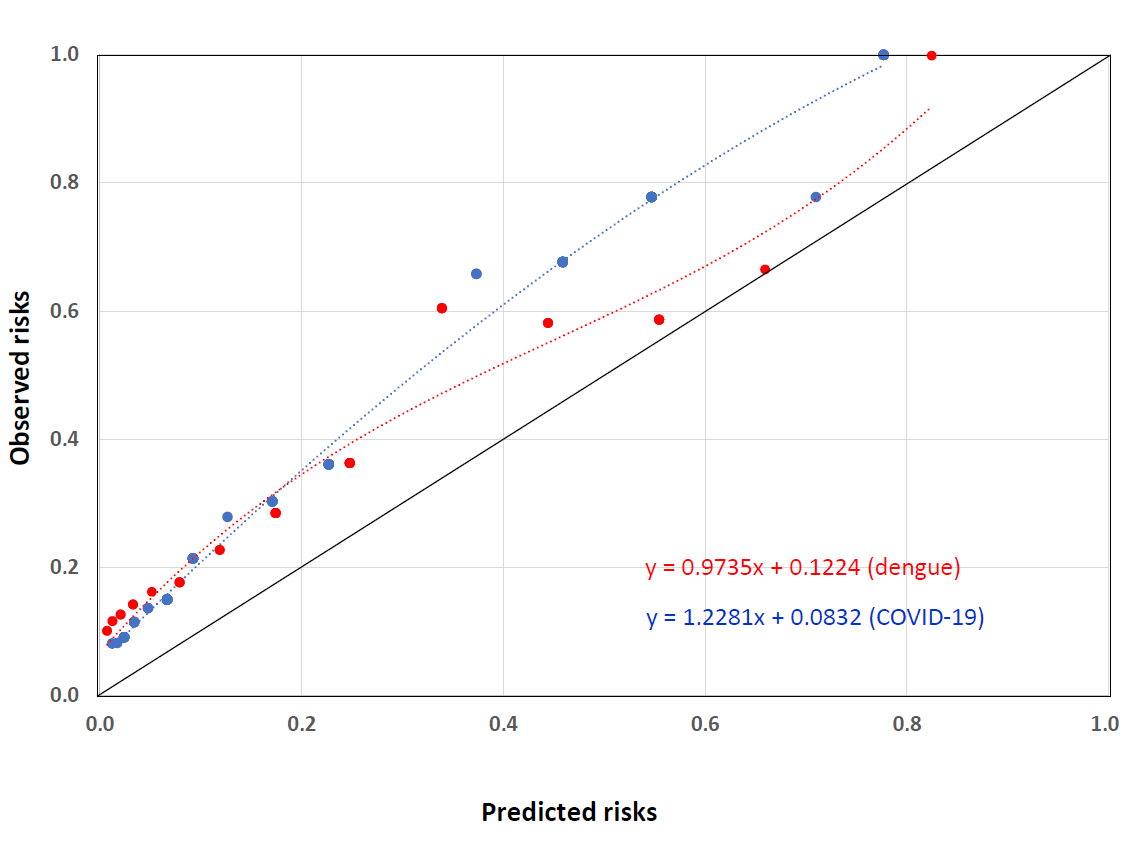

**S2 Figure. Parametric calibration plots for the COVIDENGUE scores**

Notes: Calibration plots are given for the COVIDENGUE scores before internal validation by bootstrapping. Calibrations curves display observed risks (y axis) as a function predicted risks (x axis). They are drawn using Cox nominal logistic recalibration framework. Under these conditions, perfect calibration is obtained for both an intercept equal zero and a slope equal 1 (diagonal), a slope<<1 indicates overfitting, whereas a slope>1 indicates underfitting. The panel suggests an excellent calibration for the dengue score and a relative underfitting of the COVID-19 score with a trend toward a slight miscalibration (underestimation of predicted risks) for the COVID-19 above a predicted risk of 0.25.

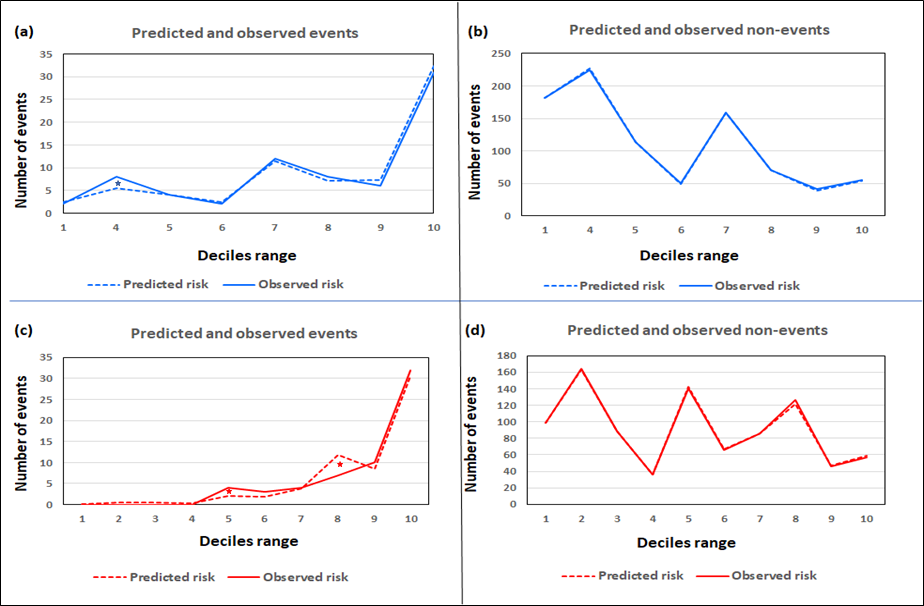

**S3 Figure. Event-based multinomial Hosmer-Lemeshow calibration plots for the COVIDENGUE scores**

Notes: Event-based calibration plots are given for the COVIDENGUE scores before internal validation by bootstrapping. Predicted and observed numbers of events (true positive/true negative) (y axis) are given for each decile of predicted risk by the Hosmer-Lemeshow chi2 test (x axis). The panel indicates non-significant areas of discrepancy (marked by stars) between observed and predicted events.

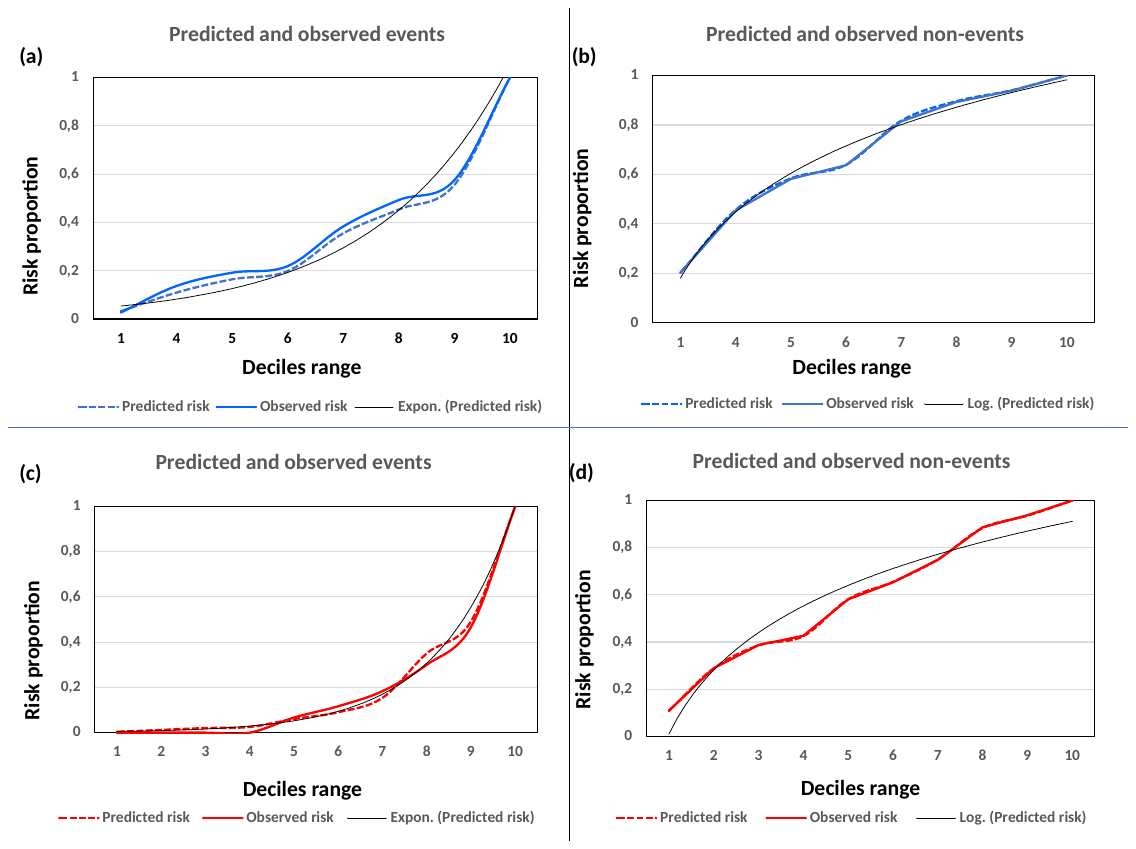

**S4 Figure. Risk-based multinomial Hosmer-Lemeshow calibration plots for the COVIDENGUE scores**

Risk-based calibration plots are given for the COVIDENGUE scores before internal validation by bootstrapping. Predicted and observed risk proportions of events (true positive/true negative) (y axis) are given for each decile of predicted risk by the Hosmer-Lemeshow chi2 test (x axis). The panel indicates a trend toward a slight miscalibration (underestimation of predicted risks) for the COVID-19 score and an excellent calibration for the dengue score
