## Supplementary material for "Differentiating COVID-19 and dengue from other febrile illnesses in co-epidemics: Development and internal validation of COVIDENGUE scores": STROBE and TRIPOD checklists

| **Section/Topic** | **Item** | **Checklist Item** | **Page** |
| --- | --- | --- | --- |
| **Title and abstract** | | | |
| Title | 1a | Indicate the study’s design with a commonly used term in the title or the abstract | 1 |
| Title | 1 | Identify the study as developing and/or validating a multivariable prediction model, the target population, and the outcome to be predicted. | 1 |
| Abstract | 1b | Provide in the abstract an informative and balanced summary of what was done and what was found | 2 |
| Abstract | 2 | Provide a summary of objectives, study design, setting, participants, sample size, predictors, outcome, statistical analysis, results, and conclusions. | 2 |
| **Introduction** | | | |
| Background, rationale and objectives | 2 | Explain the scientific background and rationale for the investigation being reported | 4 |
|  | 3a | Explain the medical context (including whether diagnostic or prognostic) and rationale for developing or validating the multivariable prediction model, including references to existing models. | 4 |
|  | 3 | State specific objectives, including any prespecified hypotheses | 4 |
|  | 3b | Specify the objectives, including whether the study describes the development or validation of the model or both. | 4 |
| **Methods** | | | |
| Study design | 4 | Present key elements of study design early in the paper | 4-5, S1* |
| Setting | 5 | Describe the setting, locations, and relevant dates, including periods of recruitment, exposure, follow-up, and data collection | 4-5, S1* |
| Source of data | 4a | Describe the study design or source of data (e.g., randomized trial, cohort, or registry data), separately for the development (DD) and validation datasets (VD), if applicable. | 4-5 (DD)  N.A (VD), S1* |
|  | 4b | Specify the key study dates, including start of accrual; end of accrual; and, if applicable, end of follow-up. | 4-5, S1* |
| Participants | 5a | Specify key elements of the study setting (e.g., primary care, secondary care, general population) including number and location of centres. | 5, S1* |
|  | 6a | *Cohort study*—Give the eligibility criteria, and the sources and methods of selection of participants. Describe methods of follow-up | 5, S1* |
|  | 6b | *Cohort study*—For matched studies, give matching criteria and number of exposed and unexposed | Not relevant |
|  | 5b | Describe eligibility criteria for participants. | 5, S1* |
|  | 5c | Give details of treatments received, if relevant. | Not relevant |
| Outcome | 7 | Clearly define all outcomes, exposures, predictors, potential confounders, and effect modifiers. Give diagnostic criteria, if applicable | 5-6, S1* |
|  | 6a | Clearly define the outcome that is predicted by the prediction model, including how and when assessed. | 5-6, S1* |
|  | 6b | Report any actions to blind assessment of the outcome to be predicted. | Not relevant |
| Variables  Predictors | 8 | For each variable of interest, give sources of data and details of methods of assessment (measurement). Describe comparability of assessment methods if there is more than one group | 5-6, S1* |
|  | 7a | Clearly define all predictors used in developing or validating the multivariable prediction model, including how and when they were measured. | 5-6, S1* |
|  | 7b | Report any actions to blind assessment of predictors for the outcome and other predictors. | Not relevant |
| Bias | 9 | Describe any efforts to address potential sources of bias (done previously, both in the body text and methodological appendix of the PLoS Negl Trop Dis article) | Ref [9] |
| Study size  Sample size | 10 | Explain how the study size was arrived at | N.A |
|  | 8 | Explain how the study size was arrived at. | N.A |
| Quantitative variables | 11 | Explain how quantitative variables were handled in the analyses. If applicable, describe which groupings were chosen and why | N.A |
| Missing data | 12c | Explain how missing data were addressed | 8, S1* |
|  | 9 | Describe how missing data were handled (e.g., complete-case analysis, single imputation, multiple imputation) with details of any imputation method. | 8, S1* |
| Statistical analysis methods | 12a | Describe all statistical methods, including those used to control for confounding | 6-8, S1* |
|  | 12b | Describe any methods used to examine subgroups and interactions | Ref [9] |
|  | 12c | *Cohort study*—If applicable, explain how loss to follow-up was addressed | N.A |
|  | 12e | Describe any sensitivity analyses | Ref [9] |
|  | 10a | Describe how predictors were handled in the analyses. | 6-8, S1* |
|  | 10b | Specify type of model, all model-building procedures (including any predictor selection), and method for internal validation. | 6-8, S1* |
|  | 10c | For validation, describe how the predictors were calculated | 7,S1* |
|  | 10d | Specify all measures used to assess model performance and, if relevant, to compare multiple models. | 7-8, S1* |
|  | 10e | Describe any model updating (e.g., recalibration) arising from the validation, if done. | N.A |
| Risk groups | 11 | Provide details on how risk groups were created, if done. | Not relevant |
| **Results** | | | |
| Participants  Descriptive data | 13a | Report numbers of individuals at each stage of study—eg numbers potentially eligible, examined for eligibility, confirmed eligible, included in the study, completing follow-up, and analysed | 9,  Fig. 1 |
|  | 13a | Describe the flow of participants through the study, including the number of participants with and without the outcome and, if applicable, a summary of the follow-up time. A diagram may be helpful. | 9,  Fig. 1 |
|  | 13b | Give reasons for non-participation at each stage | N.A |
|  | 13c | Consider use of a flow diagram | Fig. 1 |
|  | 14a | Give characteristics of study participants (eg demographic, clinical, social) and information on exposures and potential confounders | 9,  S1 Tab |
|  | 14b | Indicate number of participants with missing data for each variable of interest. | 10, Tab 1, S4, S5 Tab |
|  | 13b | Describe the characteristics of the participants (basic demographics, clinical features, available predictors), including the number of participants with missing data for predictors and outcome. | 10, Tab 1,  S4, S5 Tab |
|  | 14c | *Cohort study*—Summarise follow-up time (eg, average and total amount) | Not relevant |
| Outcome data | 15 | *Cohort study*—Report numbers of outcome events or summary measures over time | Tab 1, S2,S3, S4, S5 Tab |
| Main results | 16a | Give unadjusted estimates and, if applicable, confounder-adjusted estimates and their precision (eg, 95% confidence interval). Make clear which confounders were adjusted for and why they were included | 11-12, Tab , S2, S3 S4, S5 Tab |
|  | 16b | Report category boundaries when continuous variables were categorized | N.A |
|  | 16c | (If relevant, consider translating estimates of relative risk into absolute risk for a meaningful time period | N.A |
| Model development | 14a | Specify the number of participants and outcome events in each analysis. | Tab 1,  S2, S3, S4, S5, S6 Tab |
|  | 14b | If done, report the unadjusted association between each candidate predictor and outcome. | S2, S3 Tab |
| Model specification | 15a | Present the full prediction model to allow predictions for individuals (i.e., all regression coefficients, and model intercept or baseline survival at a given time point). | 12,  Tab 1 caption |
|  | 15b | Explain how to the use the prediction model. | 12 |
| Model performance | 16 | Report performance measures (with CIs) for the prediction model. | 11-12,  Fig 2,Fig 3, S1 fig |
| Other analyses | 17 | Report other analyses done—eg analyses of subgroups and interactions, and sensitivity analyses | Not relevant |
| **Discussion** | | | |
| Key results | 18 | Summarise key results with reference to study objectives | 13 |
| Limitations | 19 | Discuss limitations of the study, taking into account sources of potential bias or imprecision. Discuss both direction and magnitude of any potential bias | 13-15 |
|  | 18 | Discuss any limitations of the study (such as nonrepresentative sample, few events per predictor, missing data). | 13-15 |
| Interpretation | 20 | Give a cautious overall interpretation of results considering objectives, limitations, multiplicity of analyses, results from similar studies, and other relevant evidence | 15-16 |
|  | 19a | For validation, discuss the results with reference to performance in the development data, and any other validation data | N.A |
|  | 19b | Give an overall interpretation of the results, considering objectives, limitations, and results from similar studies, and other relevant evidence. | 15-16 |
| Generalisability | 21 | Discuss the generalisability (external validity) of the study results | 16-17 |
| Implications | 20 | Discuss the potential clinical use of the model and implications for future research. | 17-18 |
| **Other information** | | | |
| Supplementary information | 21 | Provide information about the availability of supplementary resources, such as study protocol, Web calculator, and data sets. | 8, 19 |
| Funding | 22 | Give the source of funding and the role of the funders for the present study and, if applicable, for the original study on which the present article is based | 20 |
|  | 22 | Give the source of funding and the role of the funders for the present study. | 20 |

* S1 file is a methodological appendix recapitulating all the methods.

We recommend using the TRIPOD Checklist in conjunction with the TRIPOD Explanation and Elaboration document.
